# Bias, precision and timeliness of historical (background) rate comparison methods for vaccine safety monitoring: an empirical multi-database analysis

**DOI:** 10.1101/2021.07.10.21258463

**Authors:** Xintong Li, Lana YH Lai, Anna Ostropolets, Faaizah Arshad, Eng Hooi Tan, Paula Casajust, Thamir M Alshammari, Talita Duarte-Salles, Evan P Minty, Carlos Areia, Nicole Pratt, Patrick B Ryan, George Hripcsak, Marc A Suchard, Martijn J Schuemie, Daniel Prieto-Alhambra

**Author notes:** **Corresponding Author: Martijn Schuemie**, Observational Health Data Analytics, Janssen R&D, Titusville, NJ, USA.

## Abstract

Using real-world data and past vaccination data, we conducted a large-scale experiment to quantify bias, precision and timeliness of different study designs to estimate historical background (expected) compared to post-vaccination (observed) rates of safety events for several vaccines. We used negative (not causally related) and positive control outcomes. The latter were synthetically generated true safety signals with incident rate ratios ranging from 1.5 to 4.

Observed vs. expected analysis using within-database historical background rates is a sensitive but unspecific method for the identification of potential vaccine safety signals. Despite good discrimination, most analyses showed a tendency to overestimate risks, with 20%-100% type 1 error, but low (0% to 20%) type 2 error in the large databases included in our study. Efforts to improve the comparability of background and post-vaccine rates, including age-sex adjustment and anchoring background rates around a visit, reduced type 1 error and improved precision but residual systematic error persisted. Additionally, empirical calibration dramatically reduced type 1 to nominal but came at the cost of increasing type 2 error.

Our study found that within-database background rate comparison is a sensitive but unspecific method to identify vaccine safety signals. The method is positively biased, with low (<=20%) type 2 error, and 20% to 100% of negative control outcomes were incorrectly identified as safety signals due to type 1 error. Age-sex adjustment and anchoring background rate estimates around a healthcare visit are useful strategies to reduce false positives, with little impact on type 2 error.

Sufficient sensitivity was reached for the identification of safety signals by month 1-2 for vaccines with quick uptake (e.g., seasonal influenza), but much later (up to month 9) for vaccines with slower uptake (e.g., varicella-zoster or papillomavirus). Finally, we reported that empirical calibration using negative control outcomes reduces type 1 error to nominal at the cost of increasing type 2 error.

## INTRODUCTION

As regulators across the world evaluate the first signals of post-marketing safety potentially associated with coronavirus disease 2019 (COVID-19) vaccines, they rely on the use of historical comparisons with so-called “background rates” for the events of interest to identify outcomes appearing more often than expected following vaccination. However, a literature gap remains on the reliability of these methods, their associated error(s), and the impact of potential strategies to mitigate them. We therefore aimed to study the bias, precision, and timeliness associated with the use of historical comparisons between post-vaccine and background rates for the identification of safety signals. We tested strategies for background rate estimation (unadjusted, age-sex adjusted, and anchored around a healthcare visit), and studied the impact of empirical calibration on type 1 and type 2 error.

## MANUSCRIPT TEXT

### BACKGROUND

One of the most common study designs in vaccine safety surveillance is the use of a cohort study with a historical comparison as a benchmark. This design allows the observed incidence of adverse events of the studied vaccine following immunization (AEFI) to be compared with the expected incidence of AEFI projected based on historical data (1). Alleged strengths include greater statistical power to detect rare AEFIs, as well as improved timeliness in detecting potential safety signals by leveraging retrospective data for analysis. There are, however, also caveats with this study design (2). Firstly, the historical population must be similar to the vaccinated cohort to obtain comparable estimates of baseline risk. Secondly, the design is subject to various temporal confounders such as seasonality, changing trends in the detection of AEFIs, and variation in diagnostic or coding criteria over time. Thirdly, the design is highly dependent on an accurate estimation of background incidence rates of the AEFIs for comparison.

Historical rate comparison has been suggested for use in several vaccine safety guidelines, including the European Network of Centres of Pharmacoepidemiology and Pharmacovigilance (ENCePP), Council for International Organizations of Medical Sciences (CIOMS), and Good Pharmacovigilance Practices (GVP). It has also been applied extensively in various clinical domains, including the Center for Disease Control and Prevention (CDC)’s Vaccine Safety Datalink (VSD) project, which used background rates to detect safety signals for the human papillomavirus vaccine (HPV) (3), adult tetanus-diphtheria-acellular pertussis (Tdap) vaccine (4), and a broad range of paediatric vaccines (5, 6). Historical data were used in Australia to detect signals for the rotavirus vaccine (7), and in Europe to detect signals for the influenza A H1N1 vaccine (8-10). While this study design is widely implemented, there is high variability in the specifics of methods used to calculate historical rates, including selection of target populations, time-at-risk windows, observation time and study settings.

### UNCERTAINTIES AND LIMITATIONS WITH THE USE OF HISTORICAL RATE COMPARISONS FOR VACCINE SAFETY MONITORING

Several studies have acknowledged uncertainties associated with the use of background rates relating to temporal and geographical variations. In one study that applied both historical comparisons and self-controlled methods, a signal of seizure in the 2014 – 2015 flu season was detected in the latter analysis but not the former. The authors explained that one possible reason was that the historical rates used might not reflect the expected baseline rate in the absence of vaccination. A second explanation was a falsely elevated background rate because of the inclusion of events induced by a previous vaccine season. Other studies have highlighted the importance of accounting for demographic, secular and seasonal trends to appropriately interpret historical rates (5, 7). Nevertheless, the influence of such trends has not been studied systematically despite observed heterogeneity in historical incidence rates (5).

It is also essential to consider the data source since there are differences in case ascertainment. This might lead to uncertainty in background rate estimates, especially in rare events(11). In addition, there might also be differences in the use of dictionary or codes to define an AEFI. For example, the spontaneous reporting system generally uses the Medical Dictionary for Regulatory Activities (MedDRA), while in observational databases different codes are used (e.g., International Classification of Diseases (ICD), SNOMED-CT, READ) and the granularity of available coding can impact the sensitivity and specificity of phenotype algorithms.

There have been suggestions on how to mitigate some of the differences between the historical and observed populations, including stratifying by age, gender, geographical or calendar time (3,5). While these approaches may reduce some differences, the distribution of the observed population is rarely known unless the study uses the spontaneous case’s demographic characteristics (of which the cases may be identified through the adverse event spontaneous system) as a proxy of the demographic characteristics of the observed population. This could potentially lead to a bias due to the estimation misclassification in each stratum based on the reporting rate (i.e. high vs. low reporting rates).

Large databases that link medical outcomes with vaccine exposure data provide a means of assessing signals identified, as well as estimates of a true incidence of clinical events after vaccination. However, these systems can be affected by relatively small denominators (given the rarity of the event) of vaccinated subjects, and a time lag in the availability of data. Very rare events or outcomes affecting a subset of the population might still be under-powered to assess a safety concern even when the data reflect the experience of millions of individuals (9). Heterogeneity in background rates across databases and age-sex strata may also persist even after robust data harmonization using common data models (12).

### OUR EXPERIMENT: INVESTIGATING THE BIAS, PRECISION AND TIMELINESS OF HISTORICAL RATE COMPARISONS FOR VACCINE SAFETY MONITORING

We aimed to fill a gap in the existing literature by estimating the bias, precision and timeliness associated with the use of historical/background compared to post-vaccination rates of safety events using ‘real world’ (electronic health records and administrative health claims) databases from the US. Our study protocol is available in the EU PAS Register (EUPAS40259)(13), and all our analytical code is in GitHub (https://github.com/ohdsi-studies/Eumaeus).

These data were previously mapped to the OMOP common data model (14). The list of included data sources, with a brief description, is available in Supplementary Table 1.

**Table 1.** Socio-demographics of participants in the contributing data sources, and number of people contributing to the analyses of different vaccines per database

| Characteristics |  | CCAE | MDCR | MDCD | Optum EHR |
| --- | --- | --- | --- | --- | --- |
| **N** |  | 156,628,301 | 10,180,158 | 31,355,646 | 97,936,862 |
| **Gender** | FEMALE | 51.12% | 55.27% | 56.23% | 53.57% |
|  | MALE | 48.88% | 44.73% | 43.77% | 46.43% |
| **H1N1 vaccination** |  | 4,598,996 | 68,102 | 1,272,745 | 916,810 |
| **Seasonal influenza vaccination** |  | 26,049,416 | 2,073,263 | 8,874,620 | 19,272,358 |
| **Zoster vaccination (Shingrix)** |  | 885,769 | 318,601 | 73,615 | 1,248,353 |
| **HPV vaccination (Gardasil 9)** |  | 2,474,810 | 28 | 1,490,831 | 1,576,255 |
| **Visit type** | Emergency Room Visit | 96,540,106 | 18,193,226 | 88,932,340 | 88,309,754 |
|  | Inpatient Visit | 26,354,555 | 9,758,649 | 17,608,722 | 55,025,828 |
|  | Outpatient Visit | 3,458,510,610 | 692,011,670 | 1,901,593,906 | 2,000,705,685 |

\*CCAE: Commercial Claims and Encounters; MDCR: Truven Health MarketScan Medicare Supplemental; MDCD: Truven Health MarketScan Medicaid; Optum EHR: Optum® de-identified Electronic Health Record Dataset.

We used retrospective data to study the following vaccines: 1) H1N1 vaccination (Sept 2009 to May 2010), 2) different types of seasonal flu vaccination (Sept 2017 to May 2018), 3) varicella-zoster vaccination (Jan 2018 to Dec 2018), and 4) HPV 9-valent recombinant vaccine (Jan 2018 to Dec 2018). Specific RxNorm codes, follow-up periods, and cohort construction details are available in Appendix – Supplementary 2. Post-vaccination rates were obtained for the period of 1 to 9 months for H1N1 and seasonal flu, and 1 to 12 for varicella-zoster and HPV vaccines. Background (historical) rates were obtained from the general population, for the same range of months one year preceding each of these vaccines. To minimise confounding, three additional variations of background rates were estimated: 1) age-sex adjusted rates; 2) visit-anchored rates; and 3) visit and age-sex adjusted rates. In the first, background rates were stratified by age (10-year bands) and sex. In the second option, background rates were estimated using the time-at-risk following a random outpatient visit. The third combined the two above to account for differences in socio-demographics and for the impact of anchoring (similar to anchoring post-vaccination in the exposed group).

We employed negative control outcomes as a benchmark to estimate bias (15, 16). Negative controls are outcomes with no plausible causal association with any of the vaccines. As such, negative control outcomes should not be identified as a signal by a safety surveillance method, and any departure from a null effect is therefore suggestive of bias due to type 1 error. A list of negative control outcomes was pre-specified for all four vaccine groups. To identify negative control outcomes that match the severity and prevalence of suspected vaccine adverse effects, a candidate list of negative controls was generated based on similarity of prevalence and percent of diagnoses that were recorded in an inpatient setting (as a proxy for severity). Manual review of this list by three clinical experts led to a final list including a total of 93 negative control outcomes. The full list is available in Supplementary Table 3.

In addition, synthetic positive control outcomes were generated to measure type 2 error (14). Given the limited knowledge of such events and the lack of consistency in the true causal association amongst other problems [6], we computed synthetic positive controls with known (albeit in silico) causal associations with the vaccines under study [5,7]. Positive outcomes were generated by modifying negative control outcomes through injection of additional simulated occurrences of the outcome, with effect sizes equivalent to true incidence rate ratios (IRR) of 1.5, 2, and 4. With the 3 mentioned true IRR, 93 negative controls were used to construct at most 93 × 3 = 279 positive control outcomes, although no positive controls were synthesized if for the negative control the number of outcomes was smaller than 25. The hazard for these outcomes was simulated to be increased for the period 1 day after vaccination until 28 days after vaccination, with a constant hazard ratio during that time.

The estimated effect size for the association vaccine-outcome was based on IRR by dividing the observed (post-vaccine) over expected (historical) incidence rates. IRR were computed both with and without empirical calibration (17). This procedure first estimates the distribution of systematic error using the negative and positive control effect-estimates and their standard derivations, then returns calibrated p-values and confidence intervals.

*Bias* was measured using: 1) Type 1 error, based on the proportion of negative control outcomes identified as safety signals according to p-value < 0.05; 2) Type 2 error, based on how often positive control outcomes were missed (not identified) as safety signals (p>0.05); 3) Area Under the receiver-operator Curve (AUC) for the discrimination of effect size estimate between positive and negative controls; and 4) Coverage, defined by how often the true IRR was within the 95% confidence interval of the estimated IRR.

*Precision* was measured using mean precision and mean squared error (MSE). Geometric mean precision was computed as 1 / (standard error)^2, with higher precision equivalent to narrower confidence intervals. MSE was obtained from the log of estimated IRRs and the log of the true HR.

To understand the time it took the analysis method to identify a safety signal (aka *timeliness*), the follow-up (up to 12 months) occurring after each vaccine was divided into calendar months. For each month, the analyses were executed using the data accumulated until the end of that month, and bias and precision metrics were estimated.

Finally, we studied the proportion of controls for which IRR were not estimable due to lack of participants exposed to the vaccine of interest. We also considered as not estimable (and therefore did not report) results for negative control outcomes with a population-based incidence rate changing>50% over time during the study period.

## FINDINGS

### Bias and precision

A total of four large databases were included, most including all four vaccines of interest: IBM MarketScan Commercial Claims and Encounters (CCAE), IBM MarketScan Multi-state Medicaid (MDCD), IBM MarketScan Medicare Supplemental Beneficiaries (MDCR), and Optum© de-identified Electronic Health Record dataset (Optum EHR). The basic socio-demographics of participants registered in each of these databases are reported in Table 1. All data sources had a majority of women, from 51.1% in CCAE to 56.23% in MDCD. As expected, data sources with older populations (e.g. IBM MDCR) had little exposure to HPV vaccination, but high numbers of participants exposed to seasonal influenza vaccination. All four data sources contributed information based on healthcare encounters in emergency rooms, outpatient as well as inpatient settings.

Historical rate comparisons were –even in their simplest form— associated with low type 2 error (0% to 10%), but led to type 1 errors ranging between 30% (HPV in MDCD) and 100% (H1N1 and seasonal flu in Optum EHR). Adjustment for age and sex reduced type 1 error in some but not all scenarios, and had limited impact on type 2 error (maximum 20% in all the conducted analyses). However, age and sex adjusted comparisons were still prone to type 1 error, with most (12/13) analyses still incorrectly identifying >=40% negative controls as potential safety signals. Anchoring the estimation of background rates around a healthcare visit helped reduce type 1 error in some scenarios (e.g., H1N1 in Optum EHR went from 100% to 50%), but increased it in others (e.g. H1N1 in CCAE increased from 50% in the unadjusted to 80% in the anchored analysis). In addition, anchoring increased type 2 error in most of our analyses, although none exceeded 20% in any of the analyses. Finally, the analyses combining anchoring and age-sex adjustment led to observable reductions in type 1 error (e.g. from 70% to 30% for HPV in CCAE), with negligible increases in type 2 error in most instances (e.g. from 10% to 20% for HPV in MDCD). Detailed results for unadjusted, age-sex adjusted, and anchoring scenarios are demonstrated in Figure 1.

**Figure 1.**
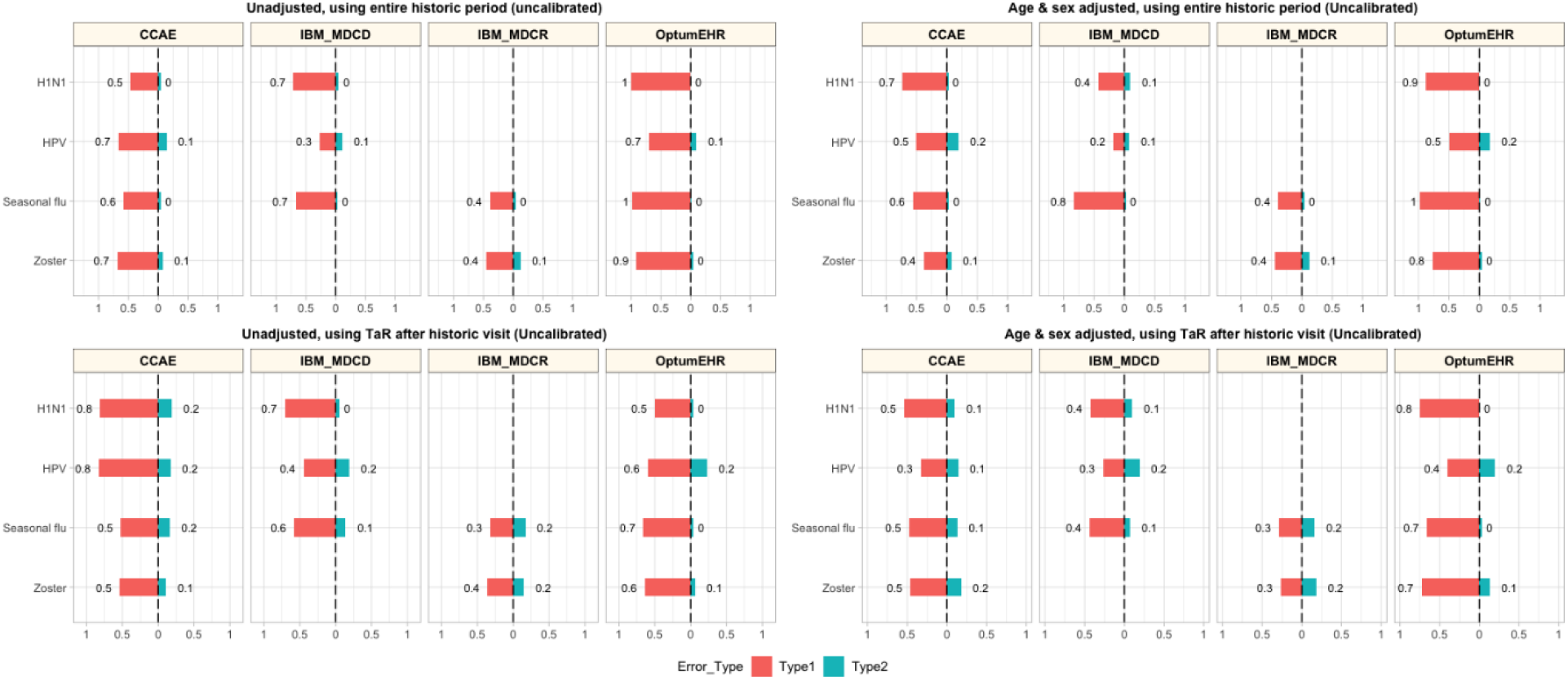
Type 1 and Type 2 error in unadjusted, age-sex adjusted, and anchored background rate analyses. CCAE: IBM MarketScan Commercial Claims and Encounters; MDCR: IBM Health MarketScan Medicare Supplemental; MDCD: IBM Health MarketScan Multi-state Medicaid; Optum EHR: Optum© de-identified Electronic Health Record Dataset.

Historical rates comparison had overall good discrimination to distinguish true safety signals (i.e. positive control outcomes), with AUCs of 80% or over in all the analyses and databases. Age-sex adjustment and anchoring had little impact on this. Conversely, coverage was low, with many analyses failing to accurately measure and include the true effect of our negative and positive control outcomes (see Table 2). Coverage in unadjusted analyses ranged from 0 (H1N1 vaccines in Optum EHR) to 0.51 (seasonal influenza vaccine in MDCR). Age-sex adjustment and anchoring had overall a positive effect on coverage, with little or no effect on discrimination (Table 2). Precision, as measured by mean precision and MSE, varied by database and vaccine exposure as reported in Table 2. Adjustment for age and sex and anchoring improved precision in most scenarios.

**Table 2.** Discrimination (AUC), coverage, precision, and MSE in unadjusted (Unadj) as well as age-sex adjusted and anchored (Adj) analyses per vaccine and data source of interest

| Vaccine exposure | Metric | CCAE |  | MDCD |  | MDCR |  | Optum EHR |  |
| --- | --- | --- | --- | --- | --- | --- | --- | --- | --- |
|  |  | Unadj | Adj | Unadj | Adj | Unadj | Adj | Unadj | Adj |
| H1N1 | AUC | 0.91 | 0.93 | 0.76 | 0.86 | N/A | N/A | 0.95 | 0.92 |
|  | Coverage | 0.42 | 0.38 | 0.18 | 0.43 | N/A | N/A | 0 | 0.22 |
|  | Mean precision | 148.85 | 132.95 | 99.1 | 75.46 | N/A | N/A | 90.18 | 83.70 |
|  | **MSE** | 0.16 | 0.09 | 0.60 | 0.17 | N/A | N/A | 0.78 | 0.23 |
| **Seasonal influenza** | **AUC** | 0.94 | 0.93 | 0.90 | 0.91 | 0.95 | 0.92 | 0.93 | 0.95 |
|  | **Coverage** | 0.29 | 0.42 | 0.26 | 0.48 | 0.51 | 0.51 | 0.01 | 0.23 |
|  | **Mean precision** | 217.94 | 154.69 | 179.27 | 133.9 | 103.87 | 80.14 | 288.58 | 220.14 |
|  | **MSE** | 0.13 | 0.10 | 0.26 | 0.12 | 0.09 | 0.09 | 0.92 | 0.11 |
| **HPV** | **AUC** | 0.82 | 0.84 | 0.88 | 0.87 | NA | NA | 0.78 | 0.84 |
|  | **Coverage** | 0.29 | 0.62 | 0.47 | 0.55 | NA | NA | 0.28 | 0.52 |
|  | **Mean precision** | 82.44 | 61.73 | 71.99 | 59.08 | NA | NA | 59.35 | 50.87 |
|  | **MSE** | 0.39 | 0.21 | 0.4 | 0.19 | NA | NA | 0.80 | 0.25 |
| **Zoster** | **AUC** | 0.86 | 0.96 | N/A | N/A | 0.87 | 0.86 | 0.84 | 0.88 |
|  | **Coverage** | 0.21 | 0.57 | N/A | N/A | 0.39 | 0.59 | 0.08 | 0.20 |
|  | **Mean precision** | 98.16 | 89.12 | N/A | N/A | 66.08 | 57.56 | 102.59 | 95.55 |
|  | **MSE** | 0.49 | 0.21 | N/A | N/A | 0.20 | 0.18 | 1.32 | 0.28 |

### The effect of empirical calibration

Empirical calibration reduced type 1 error substantially, but increased type 2 error in all the tested scenarios (see Figure 2). In addition to this, calibration improved coverage without impacting AUC, and decreased precision in most scenarios (Table 3).

**Figure 2.**
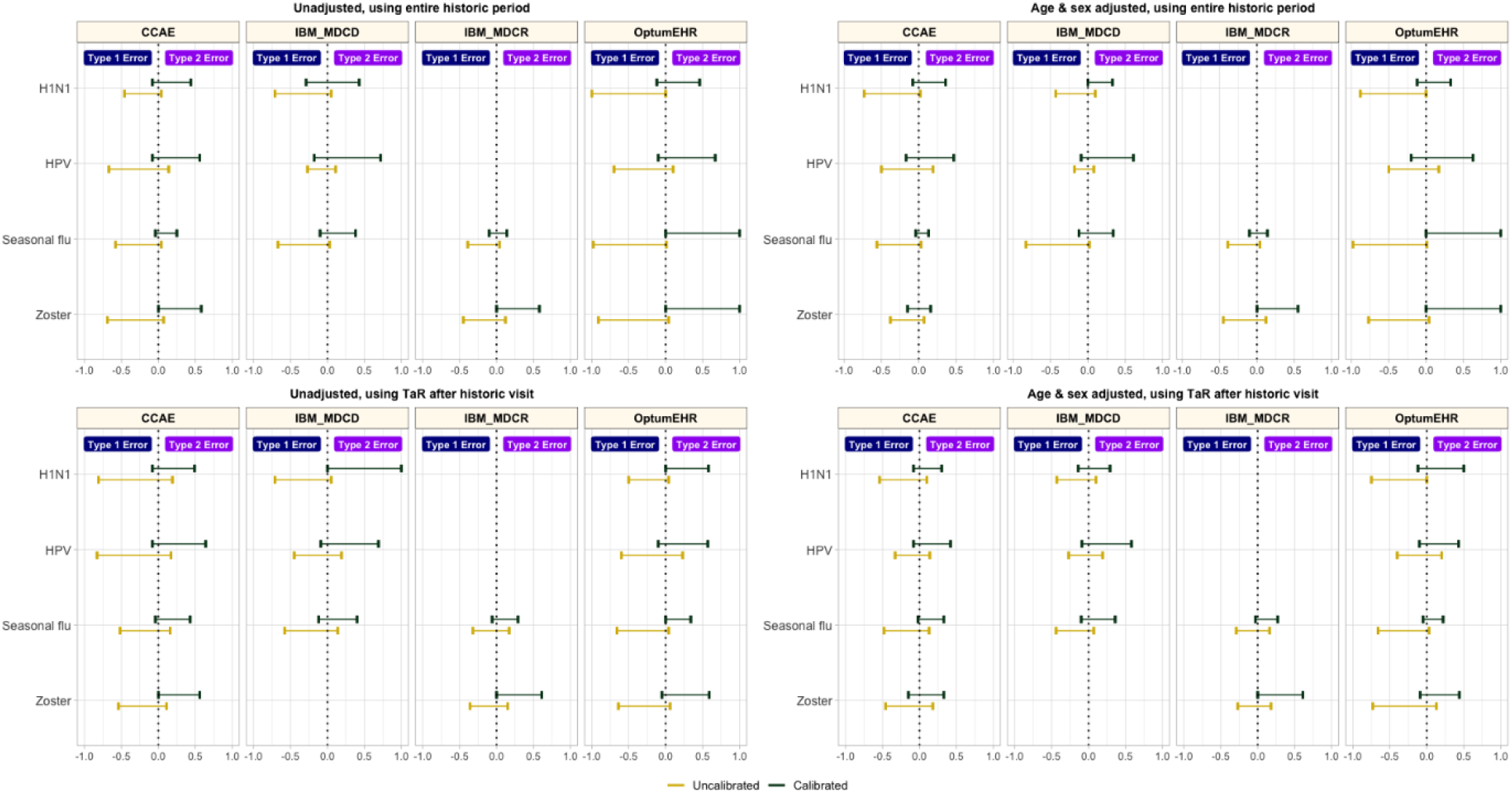
Type 1 and type 2 error before vs. after empirical calibration. *CCAE: IBM MarketScan Commercial Claims and Encounters; MDCR: IBM Health MarketScan Medicare Supplemental; MDCD: IBM Health MarketScan Multi-state Medicaid; Optum EHR: Optum© de-identified Electronic Health Record Dataset.

**Table 3.**
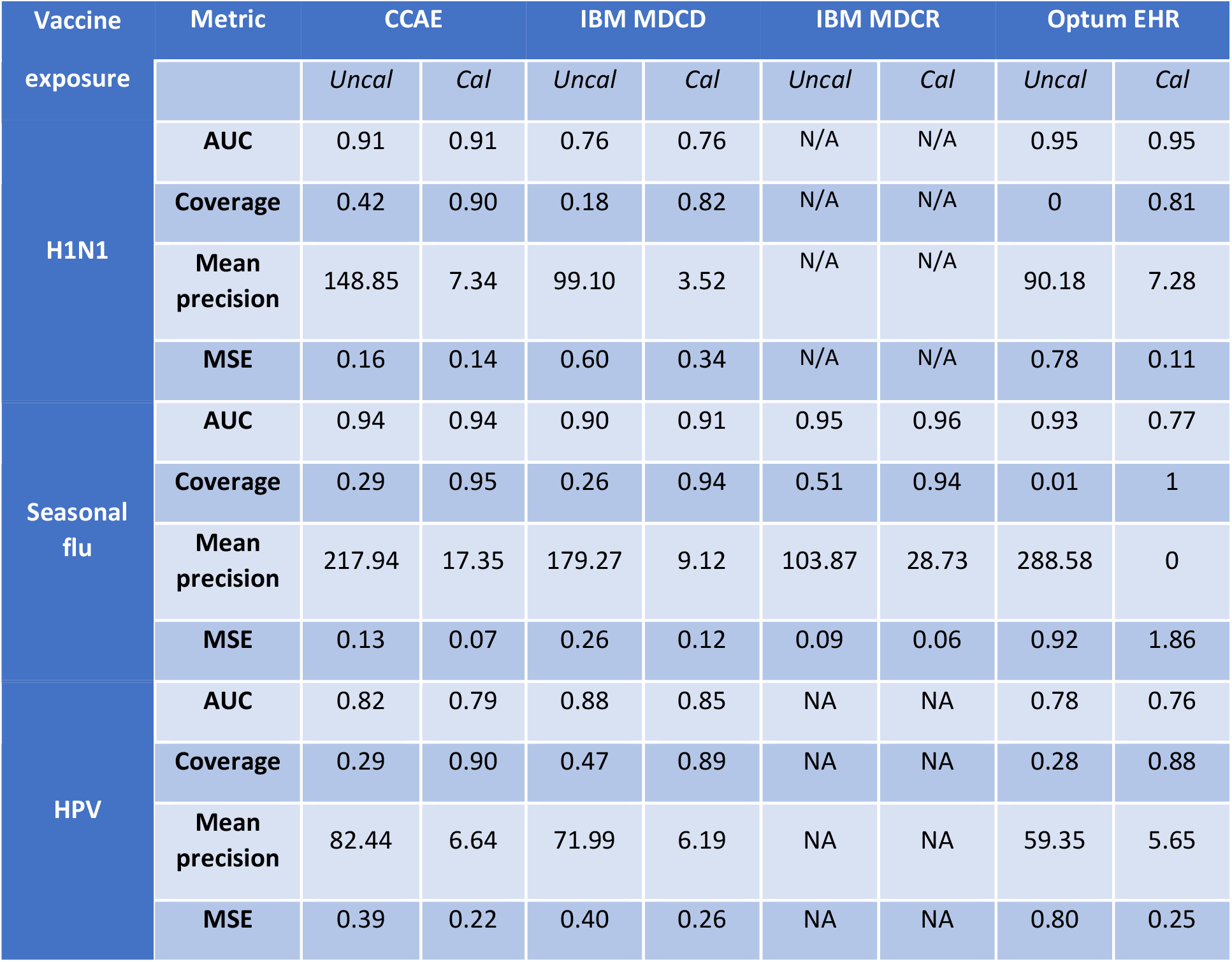
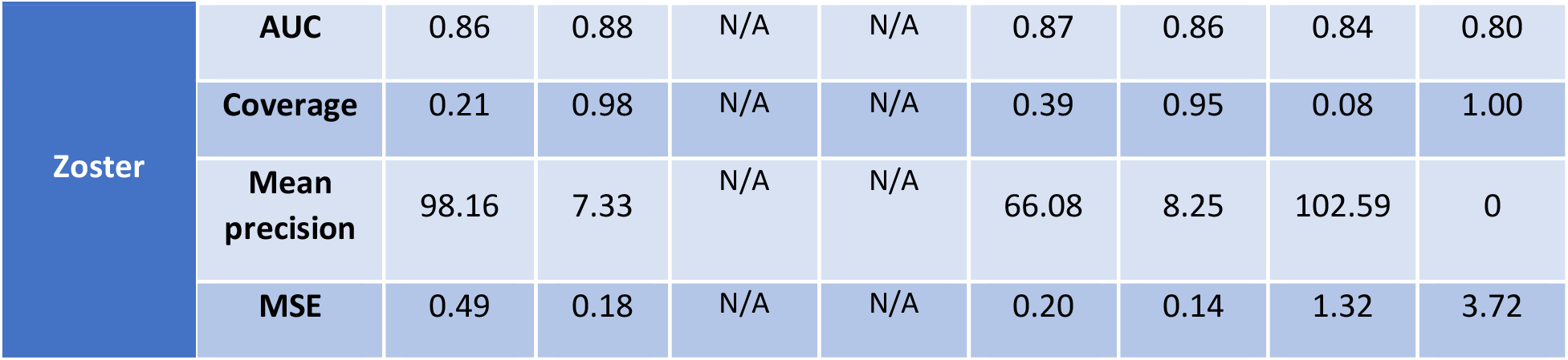
Discrimination (AUC), coverage, precision, and MSE in unadjusted analyses per vaccine and data source of interest before (Uncal) and after calibration (Cal)

### Timeliness

Most observed associations were unstable in the first few months of study, and stabilised around the true effect size in the first 2-3 months after campaign initiation for vaccines with rapid uptake like H1N1 or seasonal influenza. This stability was, however, not seen until much later, and sometimes not seen at all in the 12-month study period for vaccines with slower uptake like HPV or varicella-zoster. This is depicted in Figure 3 using data from CCAE as an illustrative example, and for all other databases in Supplementary Figures 1 to 3.

**Figure 3.**
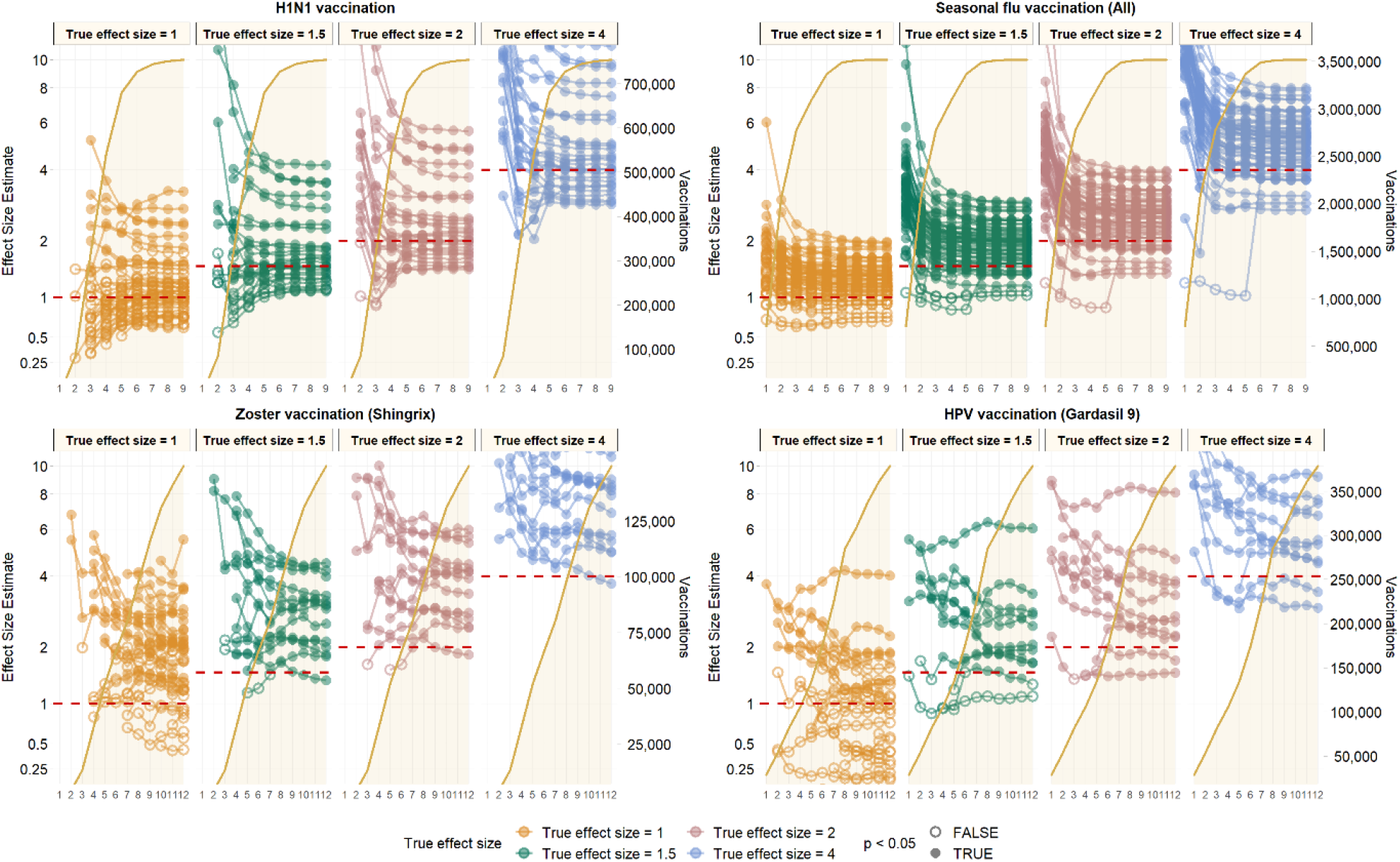
Observed effect size for negative control outcomes (true effect size = 1) and positive control outcomes (true effect size = 1.5, 2 and 4) [left Y axis] and vaccine uptake [right Y axis and shaded orange area] over time in months [X axis] based on analyses of CCAE data.

## DISCUSSION

### Key results

Our study found that unadjusted background rates comparison had low type 2 error of <10% in all analyses but unacceptably high type 1 error, up to 100% in some scenarios. The method is positively biased and uncalibrated estimates and p-values cannot be interpreted as intended; while it may be encouraging that most positive effects can be identified at a decision threshold of p<0.05, this threshold will also yield a substantial proportion of false positive findings. Age-sex adjustment and anchoring background rate estimation around a healthcare visit were useful strategies to reduce type 1 error to around 50%, while maintaining sensitivity. Empirical calibration led to restoration of type 1 error to nominal but correction for positive bias necessitates increasing type 2 error. In terms of timeliness, background rate comparisons were sensitive methods for the early identification of potential safety signals. However, most associations were exaggerated and unstable in the first few months of vaccination campaign. Vaccines with higher uptake, such as H1N1 or seasonal flu, were associated with earlier identification of safety outcomes after launch in the analyses of vaccines with rapid uptake like H1N1 or seasonal influenza.

Previous studies have shown that background incidence rates of AESI vary between age and sex (9). For example, the incidence of Bell’s palsy in adults aged over 65 years is 4 times that in paediatric population in the UK; whereas the risk of optic neuritis is higher in females than males with the same age group in Sweden. Therefore, it is crucial that age and sex are adjusted for when using background incidence rates for comparison. Nonetheless, Li et al (12) found considerable heterogeneity in incidence rates of AESI within age-sex stratified subgroups. This suggests that residual patient-level differences in characteristics such as comorbidities and medication use remained. Background rates comparison assumes that the background incidence in the overall population is similar to the vaccinated population. This assumption may not be valid because of confounding by indication, where the vaccinated population has more chronic conditions than the unvaccinated population. Conversely, the healthy vaccinee effect could occur, where on average healthier patients are more likely to adhere to annual influenza vaccination (18).

### Research in context

Post-marketing surveillance is required to ensure the safety of vaccines, so that the public do not avoid getting life-saving vaccinations because of concerns that vaccine risks are not monitored, and that any potential risks do not outweigh the vaccine’s benefits. The goal of these surveillance systems is to detect safety signals in a timely manner without raising excessive false alarms. There is an implicit trade-off between sensitivity (type 2 error) and specificity (type 1 error). Claims extending from a false positive result that is suggestive of an adverse event of a vaccine, fueled by sensationalism and unbalanced reporting in the media, could have devastating consequences on public health. A classic example of harm is the link between the MMR vaccine and autism. Although the fraudulent report by Wakefield has been retracted and many subsequent studies found no association, its lasting effects can be seen in falling MMR vaccination rates below the recommend levels from the World Health Organization (19). Expert consensus alleged that this was a contributing factor in measles being declared endemic in the United Kingdom in 2008 (20) and sporadic outbreaks in the United States in recent years (21). On the other hand, missing safety signals could put patients at risk as well as dampen public confidence in vaccination. Transparency is needed when communicating vaccination results to the public. However, it is a tricky balance to put both the benefits and harms of vaccination in context. The urgency to act quickly on the basis of incomplete real-world data could lead to confusion about vaccination safety. Negative perceptions about vaccination can be deeply entrenched and difficult to address. A starting point could be to include relevant background rates to provide comparison to other scenarios. As reported in our study, age and sex-adjusted rates are crucial to minimise false positive safety signals. Another form of communication could be using infographics to weigh harms versus benefits, illustrating the differential risks in various age groups as was shown by researchers from the University of Cambridge who contrasted the prevention of ICU admissions due to COVID-19 against the risk of blood clots due to the vaccine in specific age groups (22).

### Strengths and limitations

The strength of this study lies in the implementation of a harmonised protocol across multiple databases, which allows us to compare the findings across different healthcare systems. The use of a common data model allows the experiment to be replicated in future databases while maintaining patient privacy as patient-level data will not be shared outside of each institution. Use of real negative and synthetic positive control outcomes provides an independent estimate of residual bias in the study design and data source. The fully specified study protocol was published before analysis began and dissemination of the results did not depend on estimated effects, thus avoiding publication bias. All codes used to define the cohort, exposures, and outcomes as well as analytical code are made open source to enhance transparency and reproducibility.

In secondary use of health data, misclassification of treatment and outcomes as well as missing information due to patient care outside the respective health system are unavoidable. A strength of our study design is that it can help understand the bias inherent to the secondary use of health data.

### Future research and recommendations

When using background rate comparison for post-vaccine safety surveillance, age-sex adjustment in combination with anchoring time-at-risk around an outpatient visit resulted in somewhat reduced type 1 error, without much impact on type 2 error. Residual bias, nonetheless, remained using this design, with very high levels of type 1 error observed in most analyses. Calibration is useful for reducing Type 1 error but at the expense of decreasing precision and consequently increasing type II error. Future studies using cohort and SCCS self-controlled cased series methods with empirical calibration will be evaluated.

## Data Availability

This study is part of the Evaluating Use of Methods for Adverse Event Under Surveillance (EUMAEUS) project. The protocol of this project is available at https://ohdsi-studies.github.io/Eumaeus/Protocol.html, and we publicly host the source code at (https://github.com/ohdsi-studies/Eumaeus), allowing public contribution and review, and free re-use for anyone s future research.

## FOOTNOTES

### Contributors

XL is a PhD student in the Pharmacoepidemiology group at the Centre for Statistics in Medicine (CSM) (Oxford) and an MHS. She contributed to drafting the manuscript, creating tables and figures, and reviewing relevant literature. LYL has experience in the analysis and interpretation of routine health data, and was responsible for leading the literature review of similar studies, and for the drafting of the first version of the manuscript. AO is a PhD student and MD at Columbia University; she contributed to the analysis and the drafting of the paper. FA is an Undergraduate Student and contributed to the drafting of the manuscript and reviewing the final version of the manuscript. EHT contributed to the drafting of the manuscript. MJS is the guarantor of the study, and led the data analyses. DPA is also study guarantor and led the preparation and final review of the manuscript. All co-authors reviewed, provided feedback, and finally gave approval for the submission of the current version of the manuscript.

### Ethical approval

As this research project did not involve human subject research, it was exempted from IRB approval from the participated data partners.

### Data sharing

This study is part of the Evaluating Use of Methods for Adverse Event Under Surveillance (EUMAEUS) project. The protocol of this project is available at https://ohdsi-studies.github.io/Eumaeus/Protocol.html, and we publicly host the source code at (https://github.com/ohdsi-studies/Eumaeus), allowing public contribution and review, and free re-use for anyone’s future research.

### Transparency declaration

The lead authors affirms that this manuscript is an honest, accurate, and transparent account of the study being reported; that no important aspects of the study have been omitted; and that any discrepancies from the study as planned (and, if relevant, registered) have been explained.

### Dissemination to participants and related patient and public communities

Direct dissemination to study participants is not possible. We intend to disseminate the study results to international stakeholder groups involved with vaccine safety studies.

## Supplement materials

1. Database description
2. Exposure Cohort Definitions
3. Negative controls
4. Supplementary Figures: Observed effect size for negative control outcomes and positive control outcomes in IBM MDCD, IBM MDCR, and Optum EHR.

## Appendix

1. Database description
2. Exposure Cohort Definitions
3. Negative controls
4. Supplementary Figures: Observed effect size for negative control outcomes (true effect size = 1) and positive control outcomes in IBM MDCD, IBM MDCR, and Optum EHR.

**Table 1:** Database description

| **Data source** | **Short name** | **Description** |
| --- | --- | --- |
| Optum® de-identified Electronic Health Record Dataset (Optum EHR) | **OPTUM_EHR_US** | Optum® de-identified Electronic Health Record Dataset is derived from dozens of healthcare provider organizations in the United States (that include more than 700 hospitals and 7,000 Clinics treating more than 103 million patients) receiving care in the United States. The medical record data includes clinical information, inclusive of prescriptions as prescribed and administered, lab results, vital signs, body measurements, diagnoses, procedures, and information derived from clinical Notes using Natural Language Processing (NLP). |
| IBM MarketScan Medicare Supplemental and Coordination of Benefits Database (MDCR) | **MDCR_US** | IBM MarketScan Medicare Supplemental and Coordination of Benefits Database (MDCR) represents health services of retirees in the United States with primary or Medicare supplemental coverage through privately insured fee-for-service, point-of-service, or capitated health plans. These data include adjudicated health insurance claims (e.g. inpatient, outpatient, and outpatient pharmacy). Additionally, it captures laboratory tests for a subset of the covered lives. |
| IBM MarketScan Multi-State Medicaid Database (MDCD) | **MDCD_US** | IBM MarketScan Multi-State Medicaid Database (MDCD) contains adjudicated US health insurance claims for Medicaid enrollees from multiple states and includes hospital discharge diagnoses, outpatient diagnoses and procedures, and outpatient pharmacy claims as well as ethnicity and Medicare eligibility. Members maintain their same identifier even if they leave the system for a brief period; however the dataset lacks lab data. |
| IBM MarketScan Commercial Claims and Encounters Database (CCAE) | CCAE_US | IBM MarketScan Commercial Claims and Encounters Database (CCAE) is a US employer-based private-payer administrative claims database. The data include adjudicated health insurance claims (e.g. inpatient, outpatient, and outpatient pharmacy) as well as enrollment data from large employers and health plans who provide private healthcare coverage to employees, their spouses, and dependents. Additionally, it captures laboratory tests for a subset of the covered lives. This administrative claims database includes a variety of fee-for-service, preferred provider organizations, and capitated health plans. |

### 2. Exposure Cohort Definitions

#### 2.1 H1N1 Vaccines

#### 2.1.1 Cohort Entry Events

People enter the cohort when observing any of the following:

1. drug exposures of ‘H1N1 vaccine,’ starting between September 1, 2009 and May 31, 2010. Limit cohort entry events to the earliest event per person.

#### 2.1.2 Cohort Exit

The cohort end date will be offset from index event’s start date plus 0 days.

#### 2.1.3 Cohort Eras

Entry events will be combined into cohort eras if they are within 0 days of each other.

#### 2.1.4 H1N1 vaccine

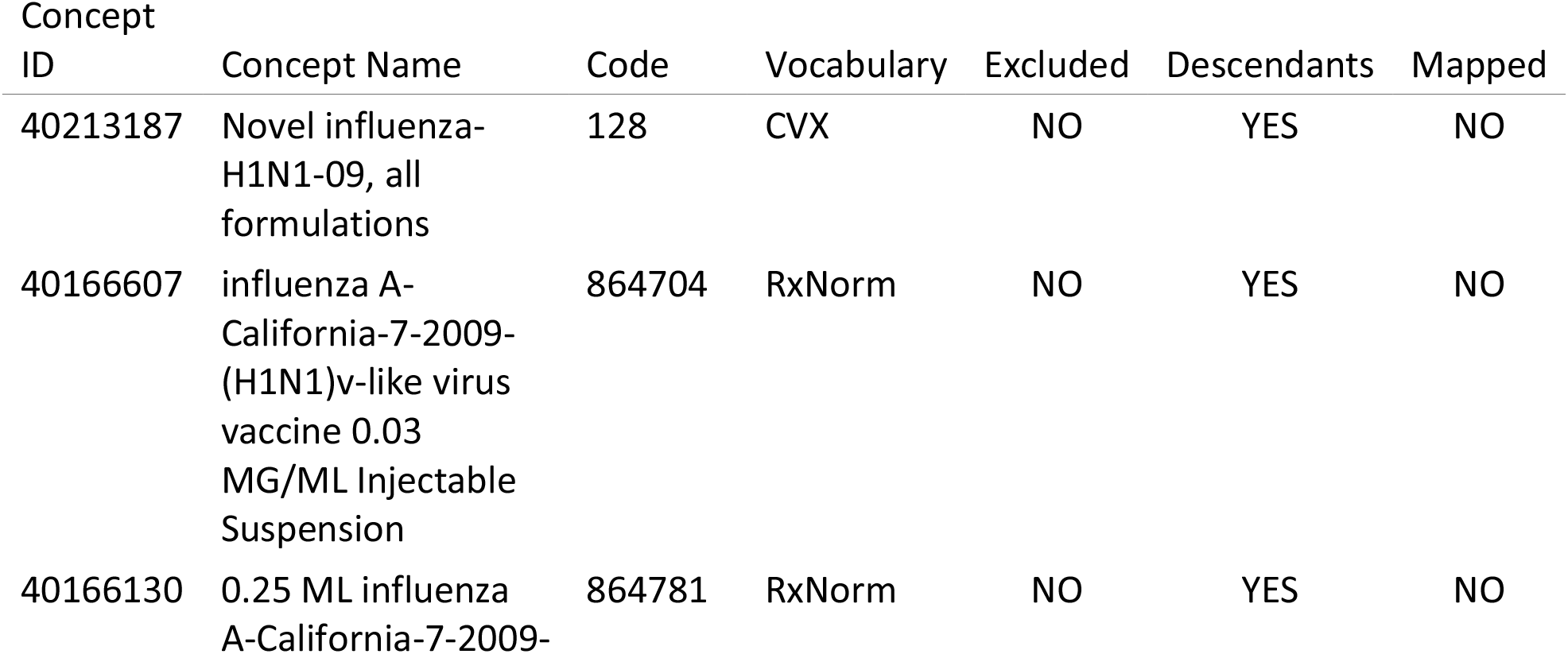
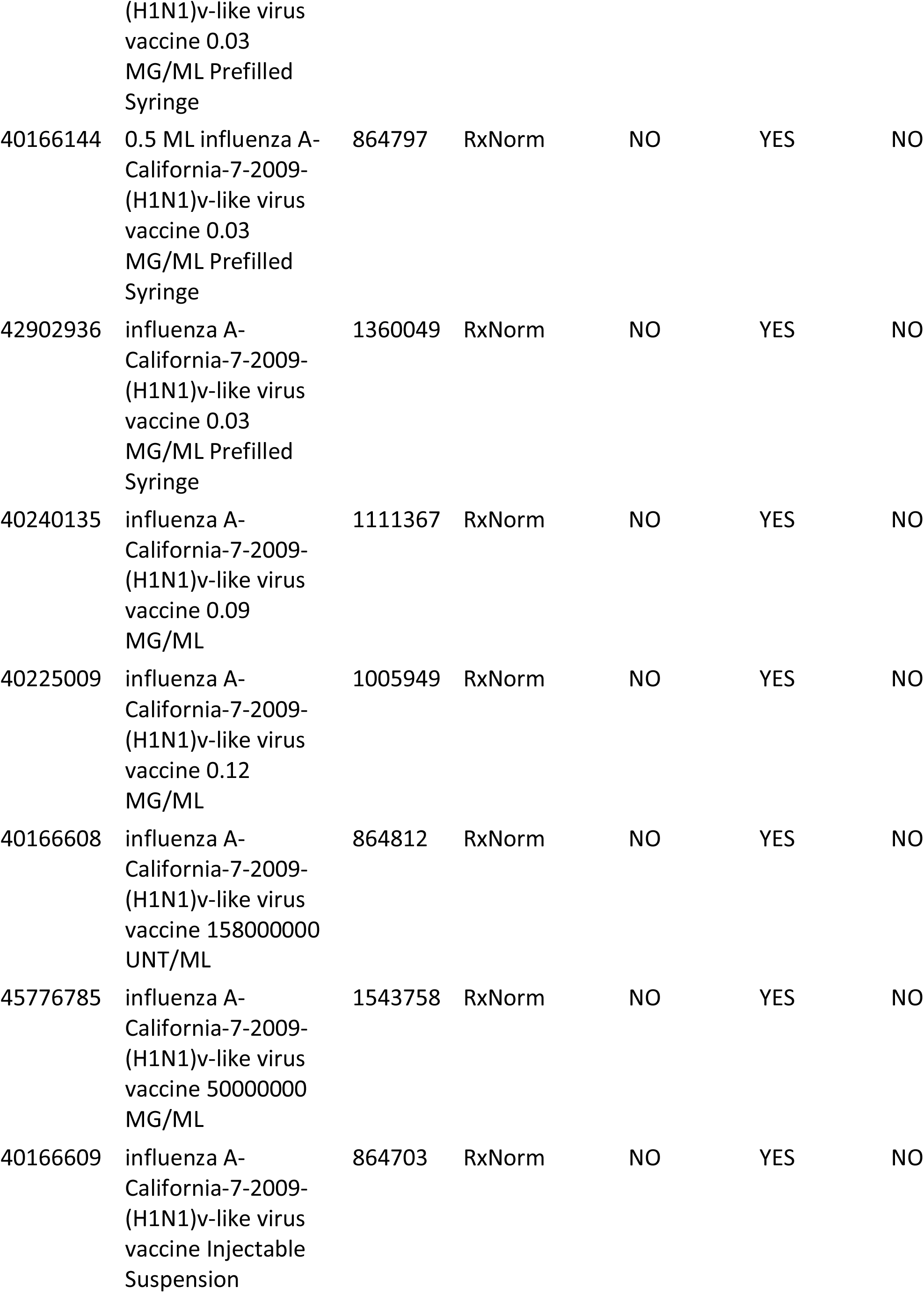
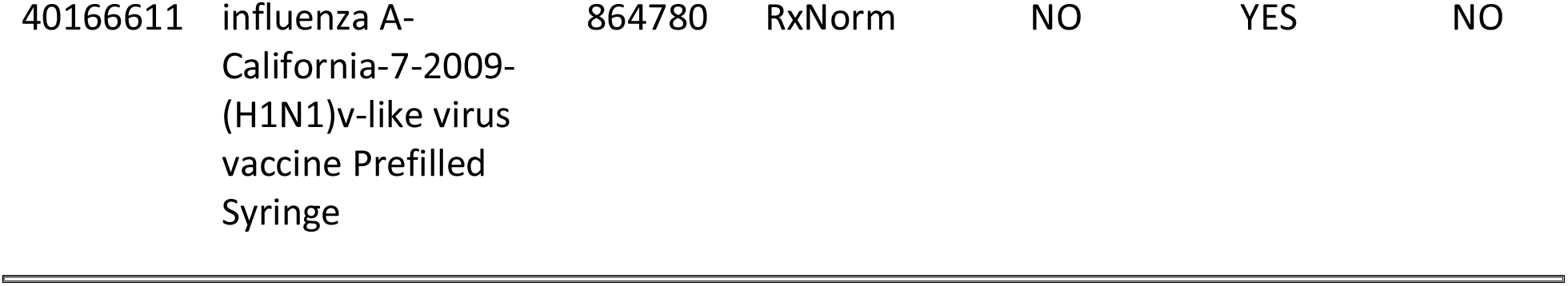

### 2.2 Seasonal Flu Vaccines (All)

#### 2.2.1 Cohort Entry Events

People enter the cohort when observing any of the following:

1. drug exposures of ‘Seasonal flu vaccine,’ starting between September 1, 2017 and May 31, 2018.

Limit cohort entry events to the earliest event per person.

#### 2.2.2 Cohort Exit

The cohort end date will be offset from index event’s start date plus 0 days.

#### 2.2.3 Cohort Eras

Entry events will be combined into cohort eras if they are within 0 days of each other.

#### 2.2.4 Seasonal flu vaccine

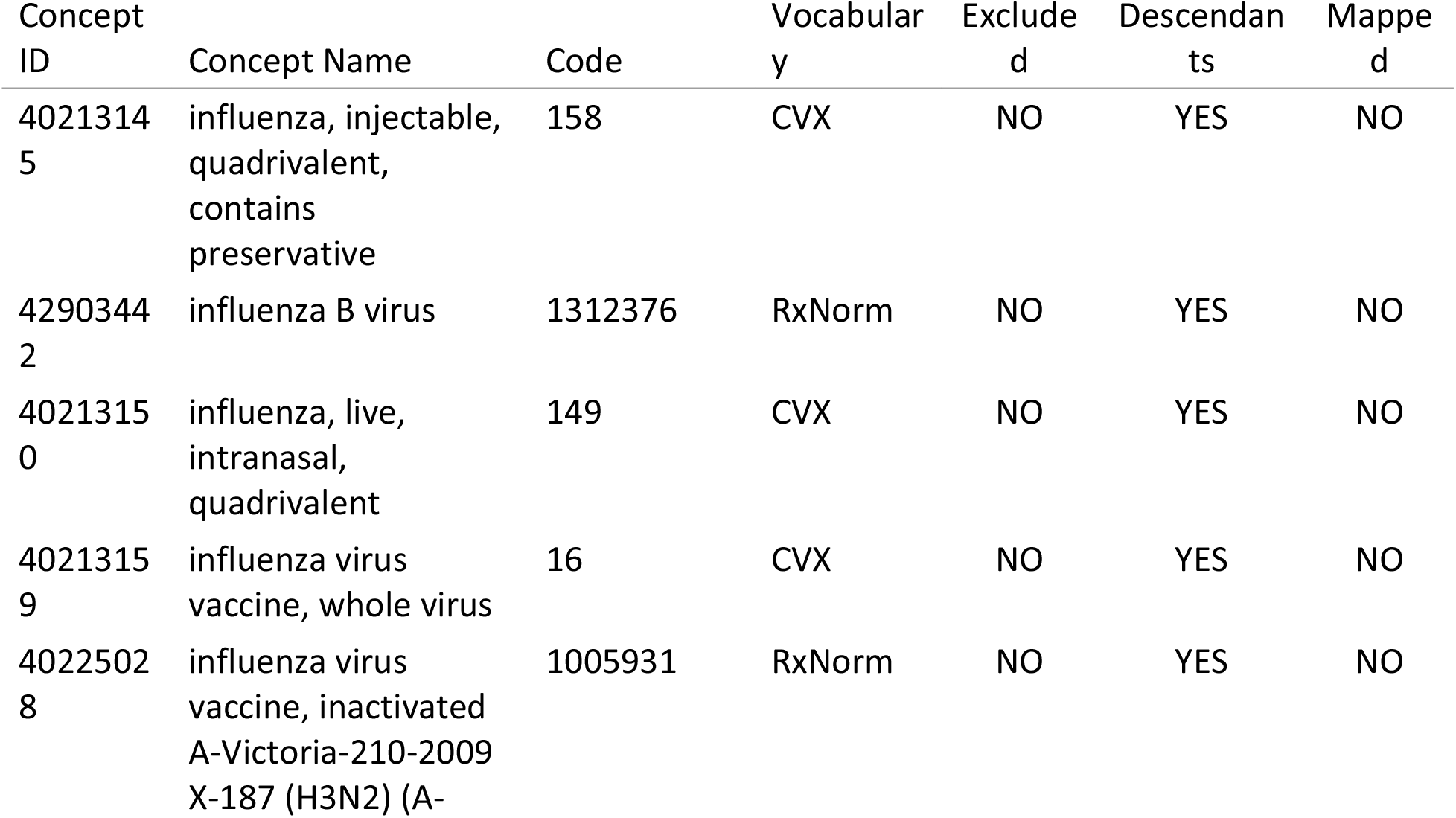
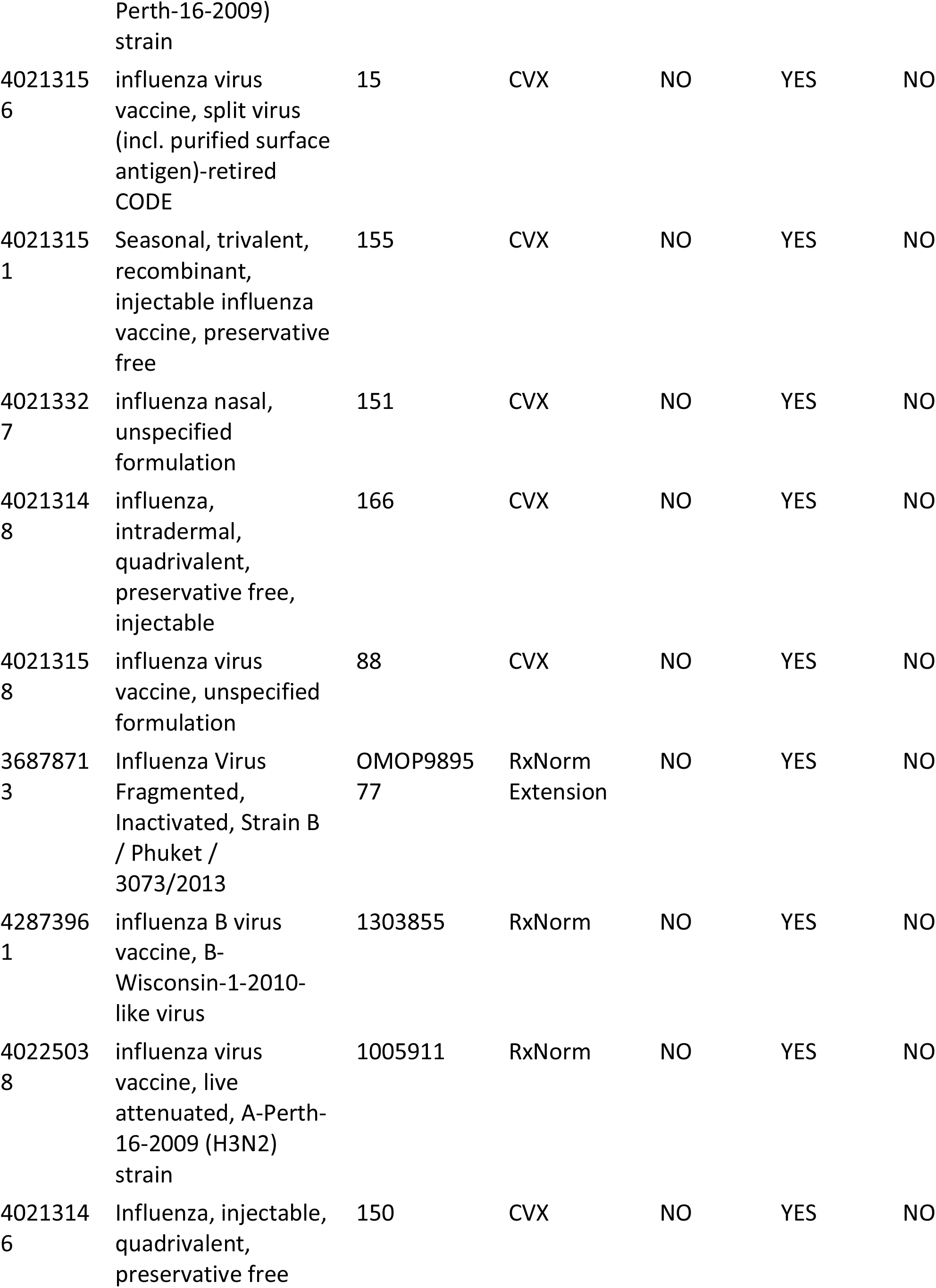
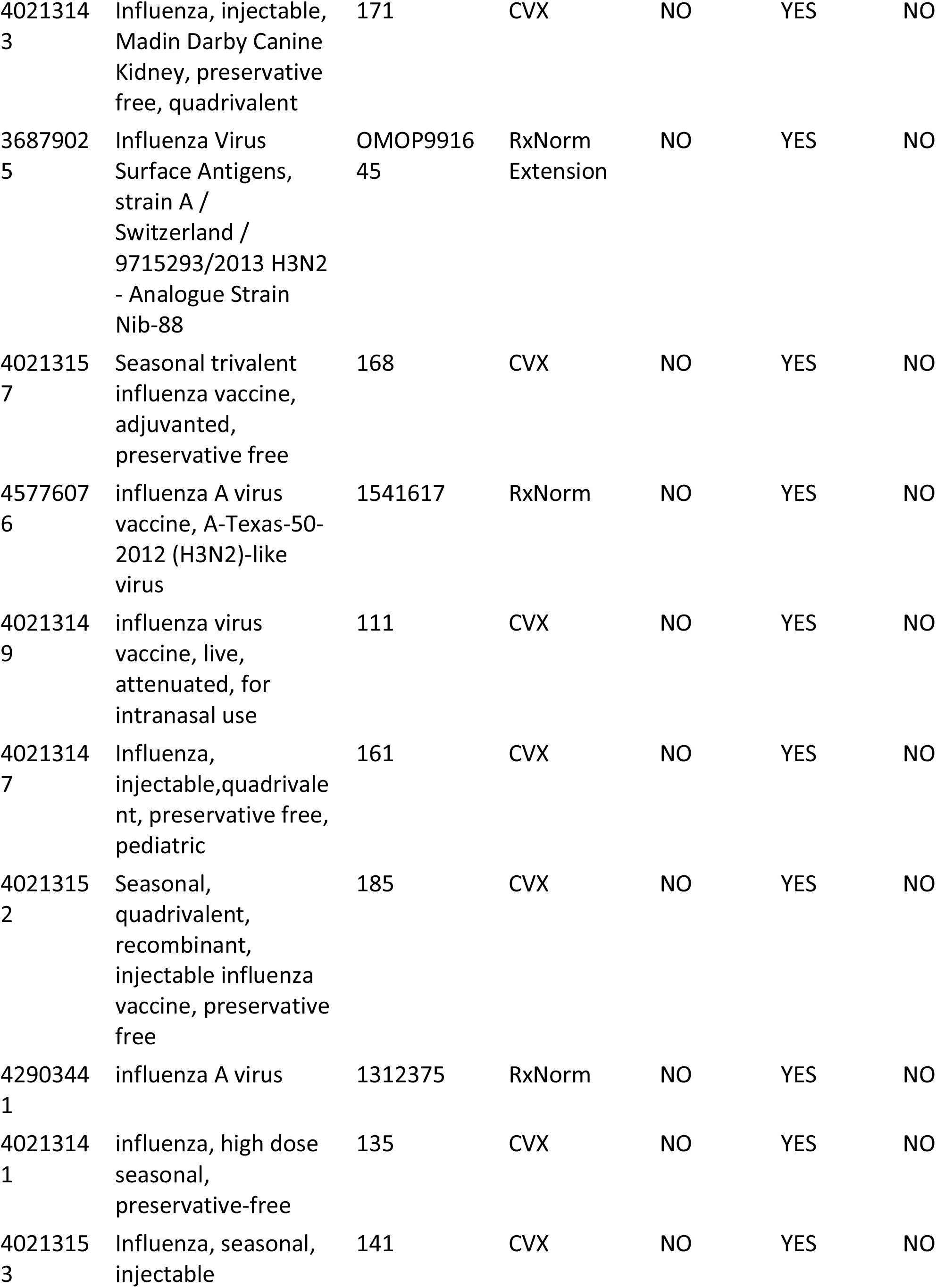
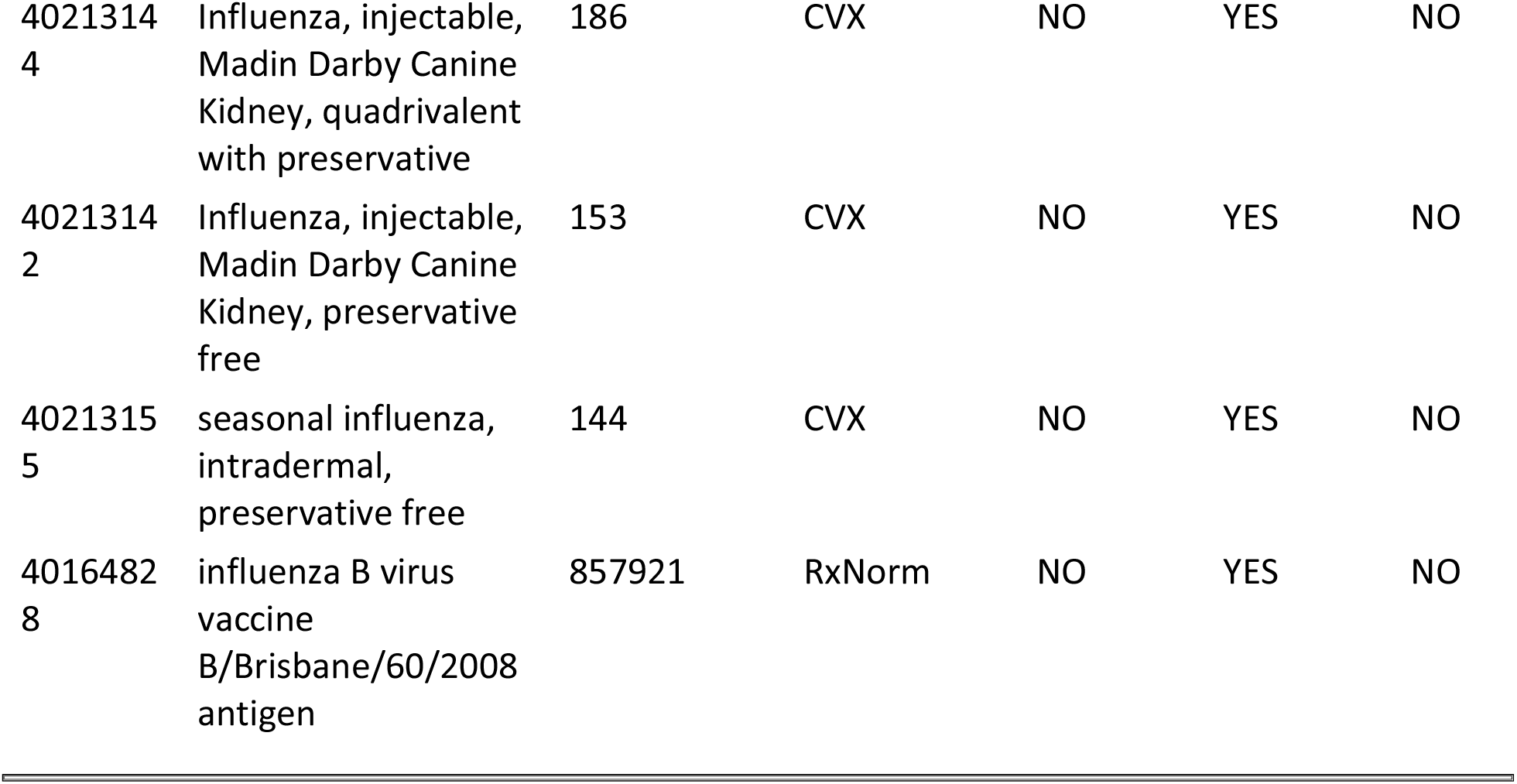

### 2.3 HPV Vaccines

#### 2.3.1 Cohort Entry Events

People enter the cohort when observing any of the following:

1. drug exposures of ‘Gardasil 9,’ starting between January 1, 2018 and December 31, 2018.

#### 2.3.2 Cohort Exit

The cohort end date will be offset from index event’s start date plus 0 days.

#### 2.3.3 Cohort Eras

Entry events will be combined into cohort eras if they are within 0 days of each other.

#### 2.3.4 Gardasil 9

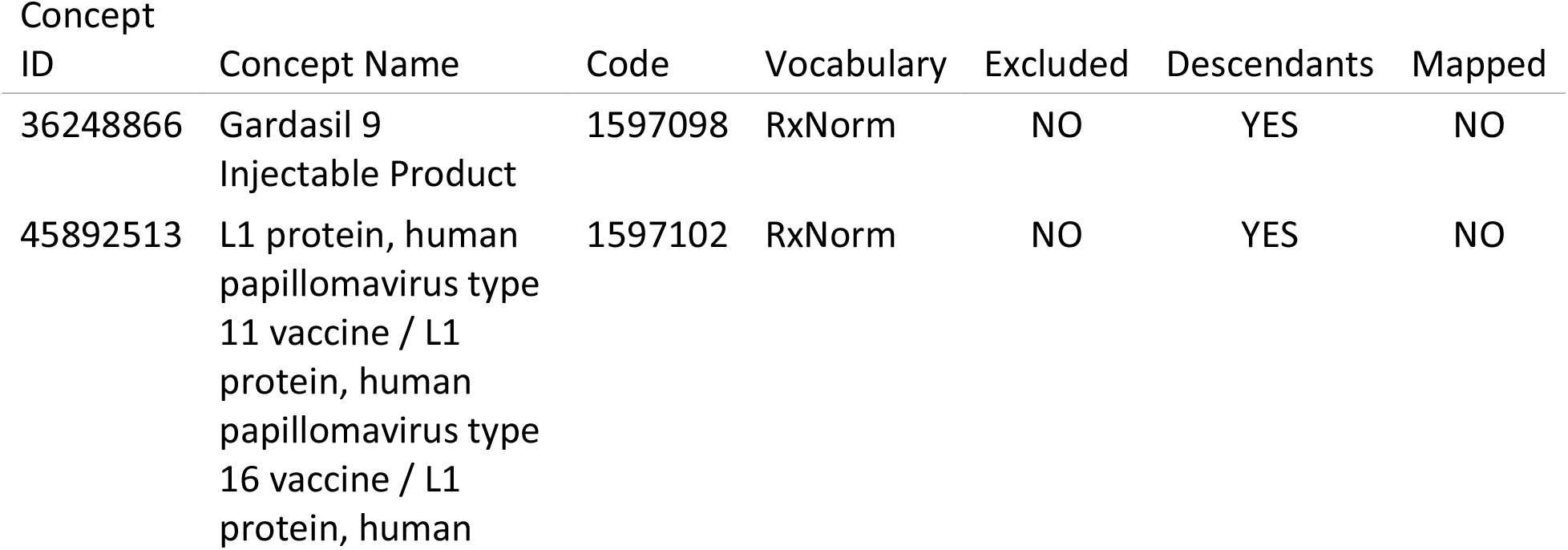
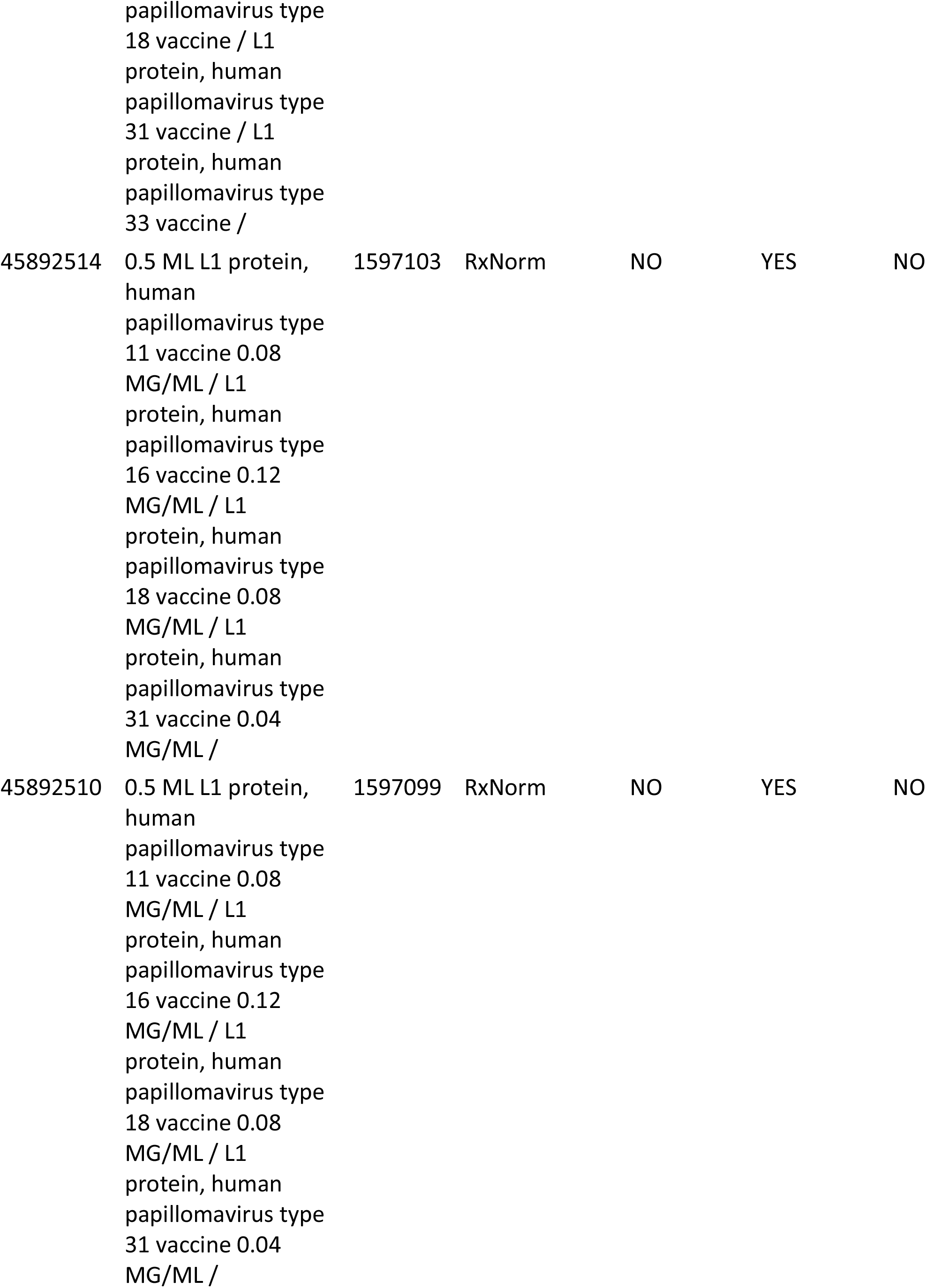
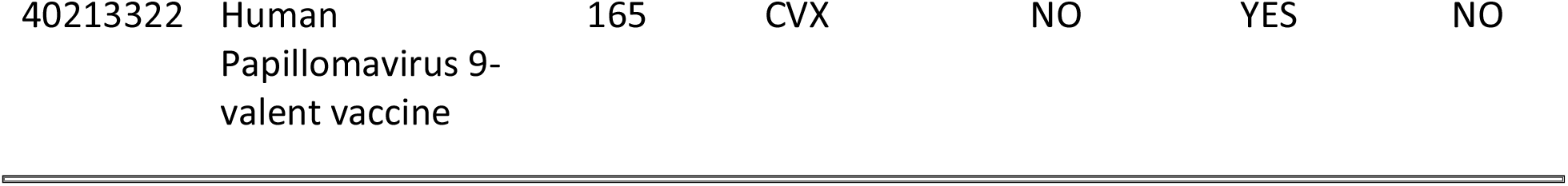

| Concept<br>ID | Concept Name | Code | Vocabulary | Excluded | Descendants | Mapped |
| --- | --- | --- | --- | --- | --- | --- |
| 36248866 | Gardasil 9<br>Injectable Product | 1597098 | RxNorm | NO | YES | NO |
| 45892513 | L1 protein, human<br>papillomavirus type<br>11 vaccine / L1<br>protein, human<br>papillomavirus type<br>16 vaccine / L1<br>protein, human papillomavirus type<br>18 vaccine / L1<br>protein, human<br>papillomavirus type<br>31 vaccine / L1<br>protein, human<br>papillomavirus type<br>33 vaccine / | 1597102 | RxNorm | NO | YES | NO |
| 45892514 | 0.5 ML L1 protein,<br>human<br>papillomavirus type<br>11 vaccine 0.08<br>MG/ML / L1<br>protein, human<br>papillomavirus type<br>16 vaccine 0.12<br>MG/ML / L1<br>protein, human<br>papillomavirus type<br>18 vaccine 0.08<br>MG/ML / L1<br>protein, human<br>papillomavirus type<br>31 vaccine 0.04<br>MG/ML / | 1597103 | RxNorm | NO | YES | NO |
| 45892510 | 0.5 ML L1 protein,<br>human<br>papillomavirus type<br>11 vaccine 0.08<br>MG/ML / L1<br>protein, human<br>papillomavirus type<br>16 vaccine 0.12<br>MG/ML / L1<br>protein, human<br>papillomavirus type<br>18 vaccine 0.08<br>MG/ML / L1<br>protein, human<br>papillomavirus type<br>31 vaccine 0.04<br>MG/ML / | 1597099 | RxNorm | NO | YES | NO |
| 40213322 | Human<br>Papillomavirus 9-valent vaccine | 165 | CVX | NO | YES | NO |

### 2.4 Zoster Vaccines

#### 2.4.1 Cohort Entry Events

People enter the cohort when observing any of the following:

1. drug exposures of ‘Shingrix,’ starting between January 1, 2018 and December 31, 2018.

#### 2.4.2 Cohort Exit

The cohort end date will be offset from index event’s start date plus 0 days.

#### 2.4.3 Cohort Eras

Entry events will be combined into cohort eras if they are within 0 days of each other.

#### 2.4.4 Shingrix

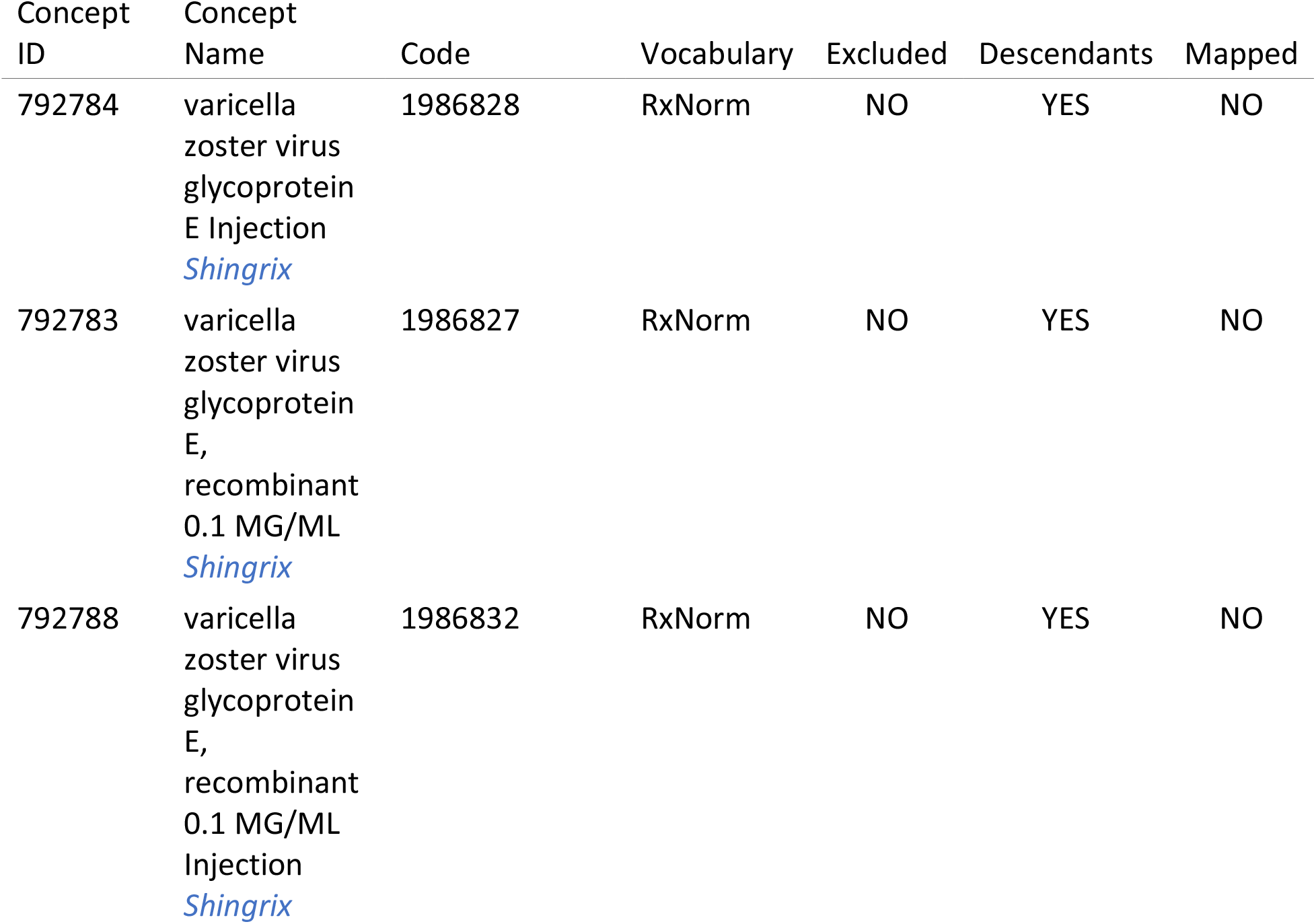
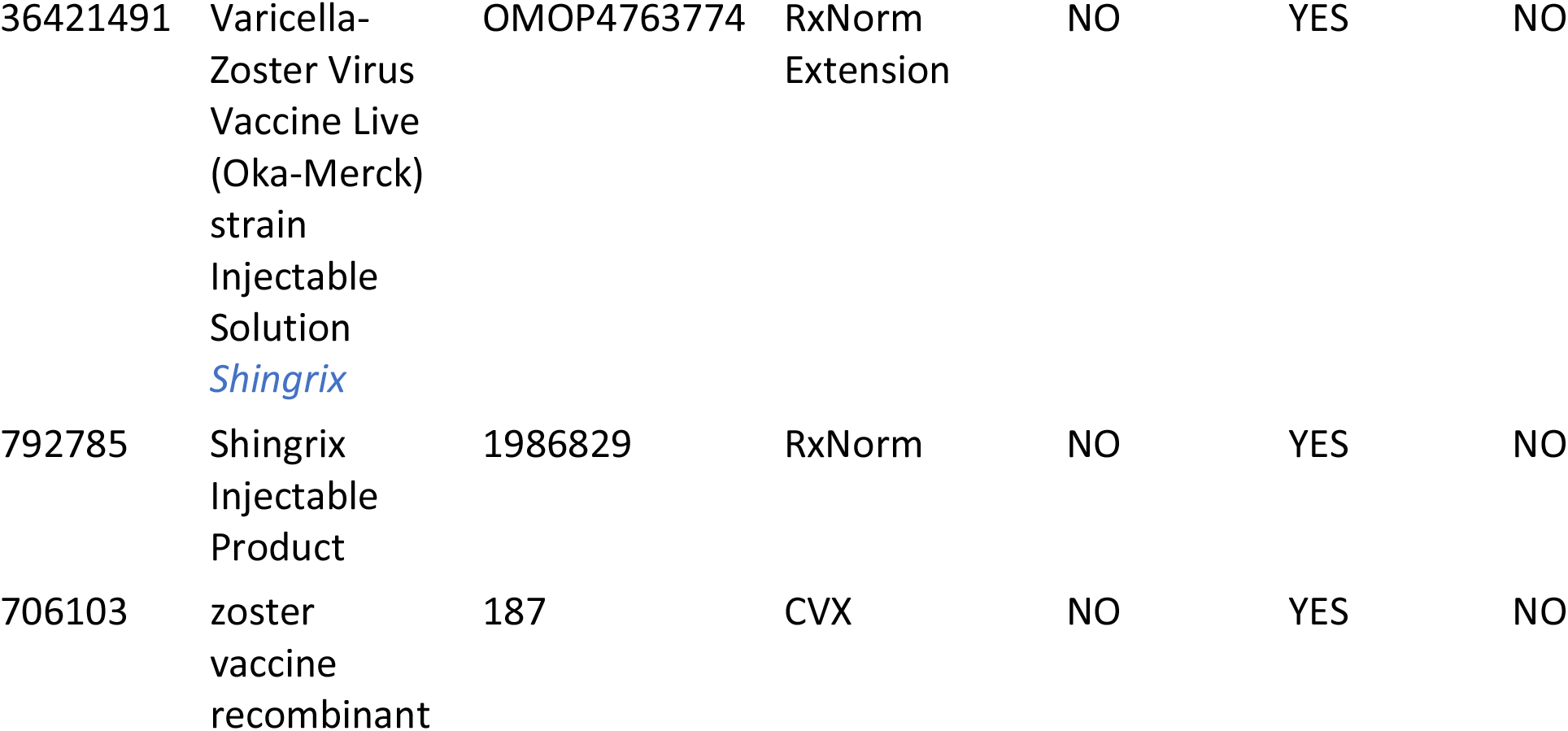

| Concept ID | Concept Name | Code | Vocabulary | Excluded | Descendants | Mapped |
| --- | --- | --- | --- | --- | --- | --- |
| 792784 | varicella<br>zoster virus<br>glycoprotein<br>E Injection<br>*Shingrix* | 1986828 | RxNorm | NO | YES | NO |
| 792783 | varicella<br>zoster virus<br>glycoprotein<br>E,<br>recombinant<br>0.1 MG/ML<br>*Shingrix* | 1986827 | RxNorm | NO | YES | NO |
| 792788 | varicella<br>zoster virus<br>glycoprotein<br>E,<br>recombinant<br>0.1 MG/ML<br>Injection<br>*Shingrix* | 1986832 | RxNorm | NO | YES | NO |
| 36421491 | Varicella-Zoster Virus Vaccine Live (Oka-Merck) strain Injectable Solution *Shingrix* | OMOP4763774 | RxNorm Extension | NO | YES | NO |
| 792785 | Shingrix Injectable Product | 1986829 | RxNorm | NO | YES | NO |
| 706103 | zoster vaccine recombinant | 187 | CVX | NO | YES | NO |

**Table 3:**
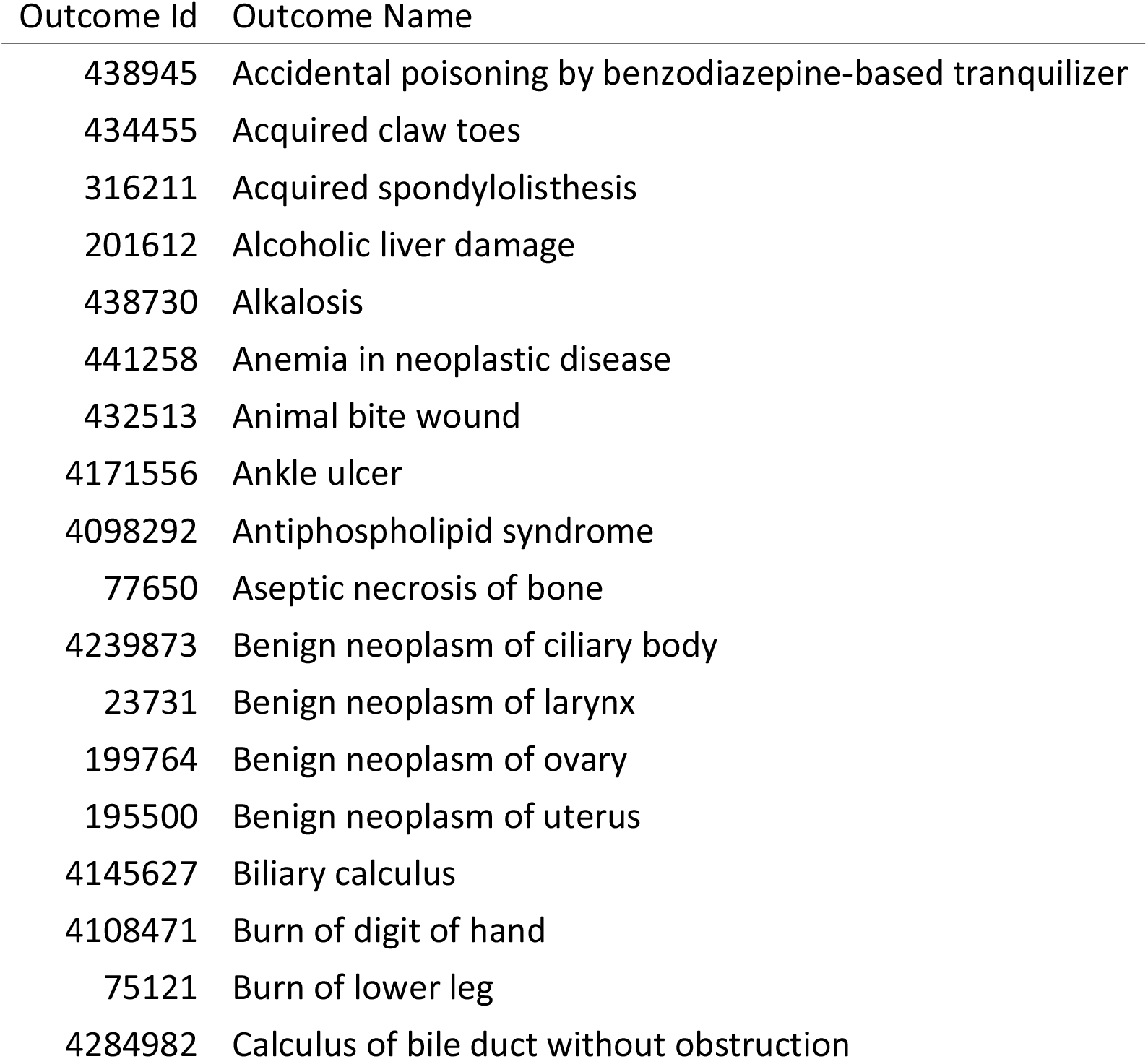
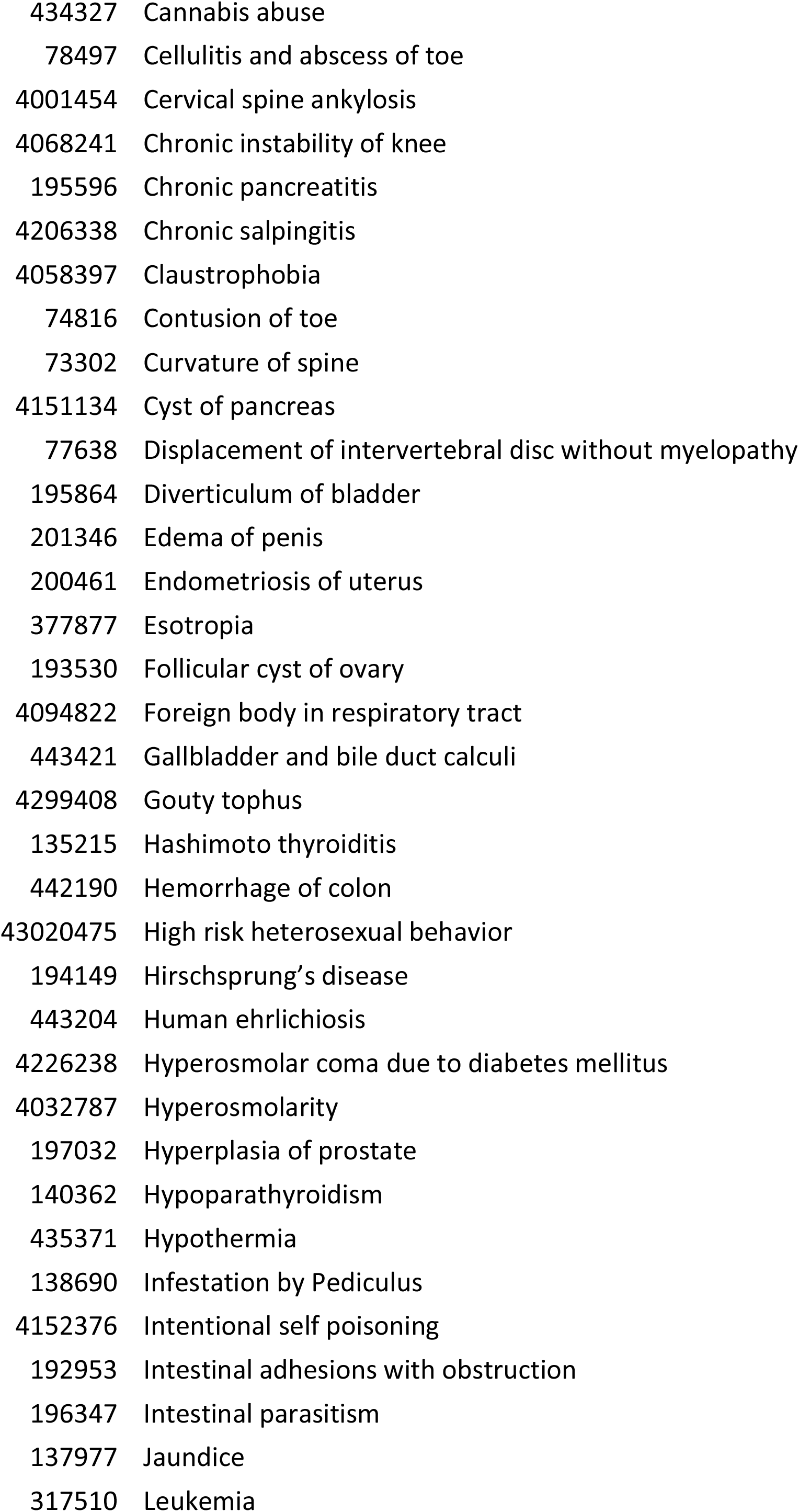
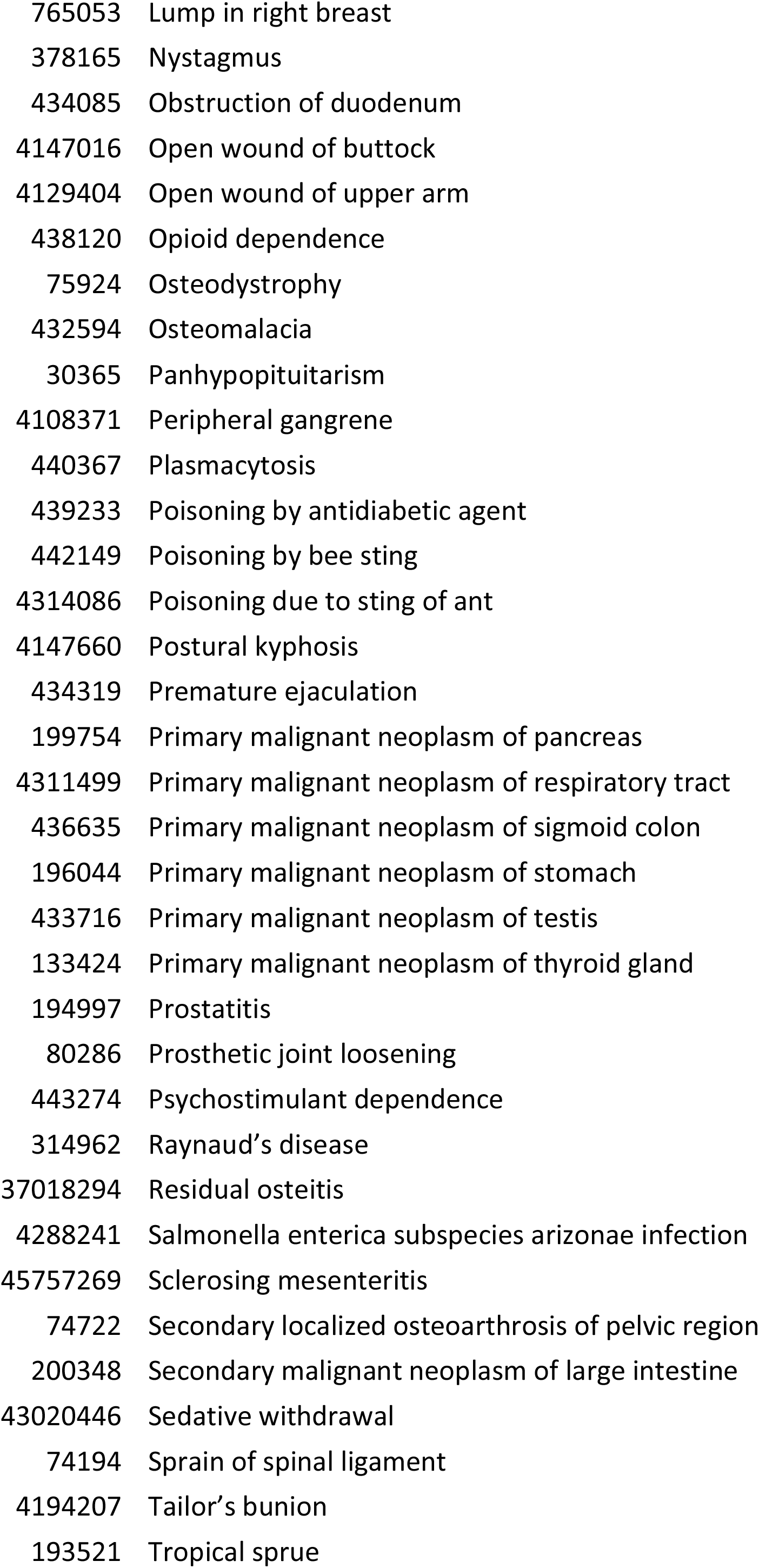
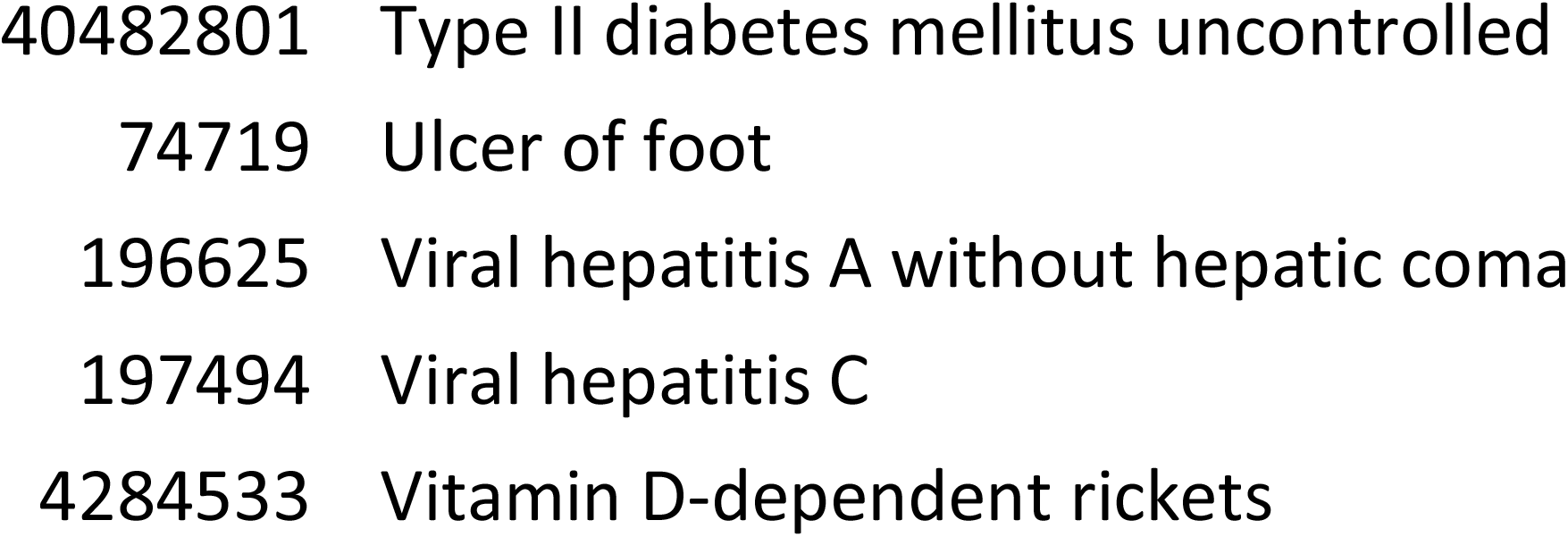
Negative control outcomes.

## Supplementary Figures

Observed effect size for negative control outcomes (true effect size = 1) and positive control outcomes (true effect size = 1.5, 2 and 4) [left Y axis] and vaccine uptake [right Y axis and shaded orange area] over time in months [X axis].

**Figure S1:**
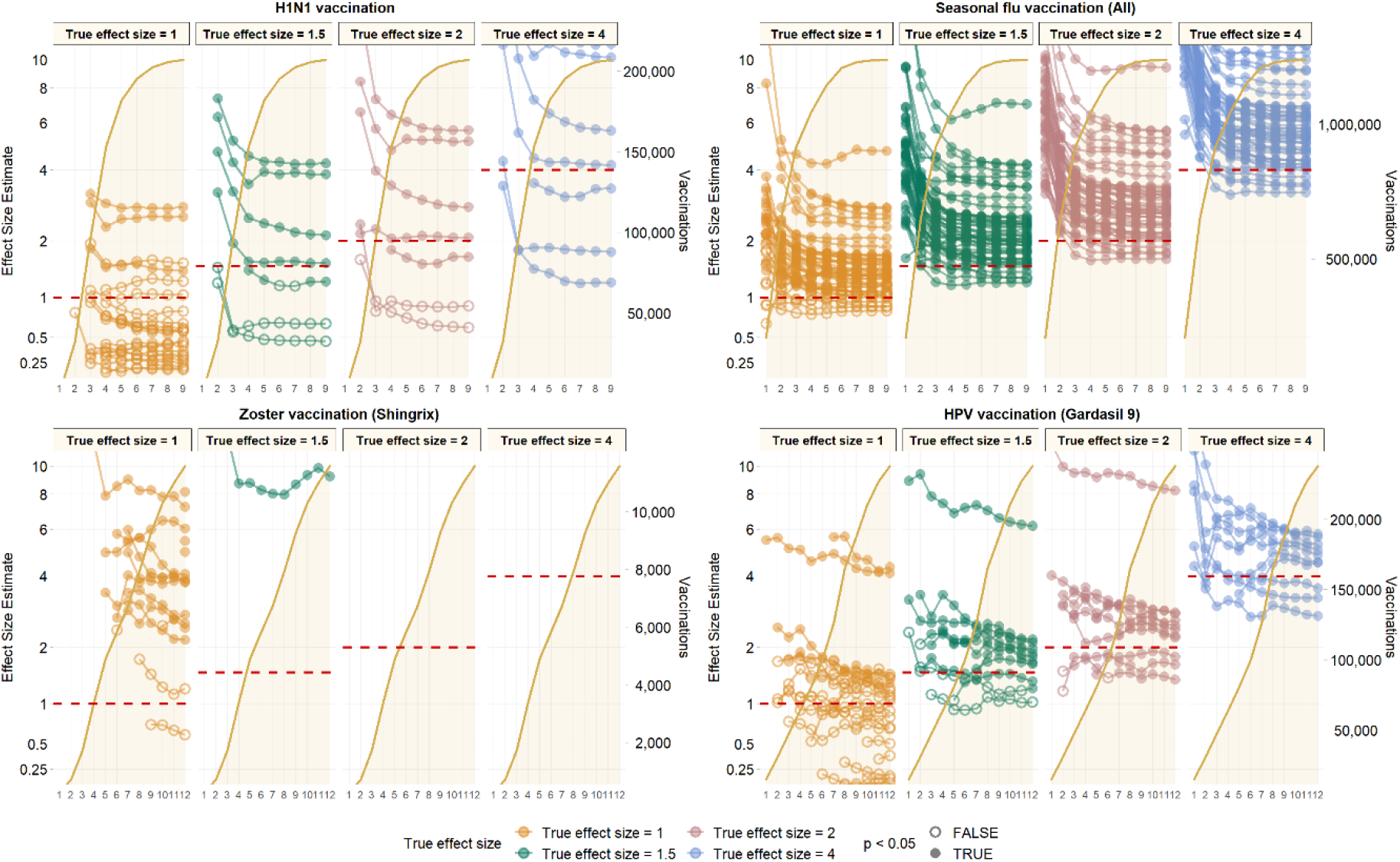
IBM MDCD (unadjusted)

**Figure S2:**
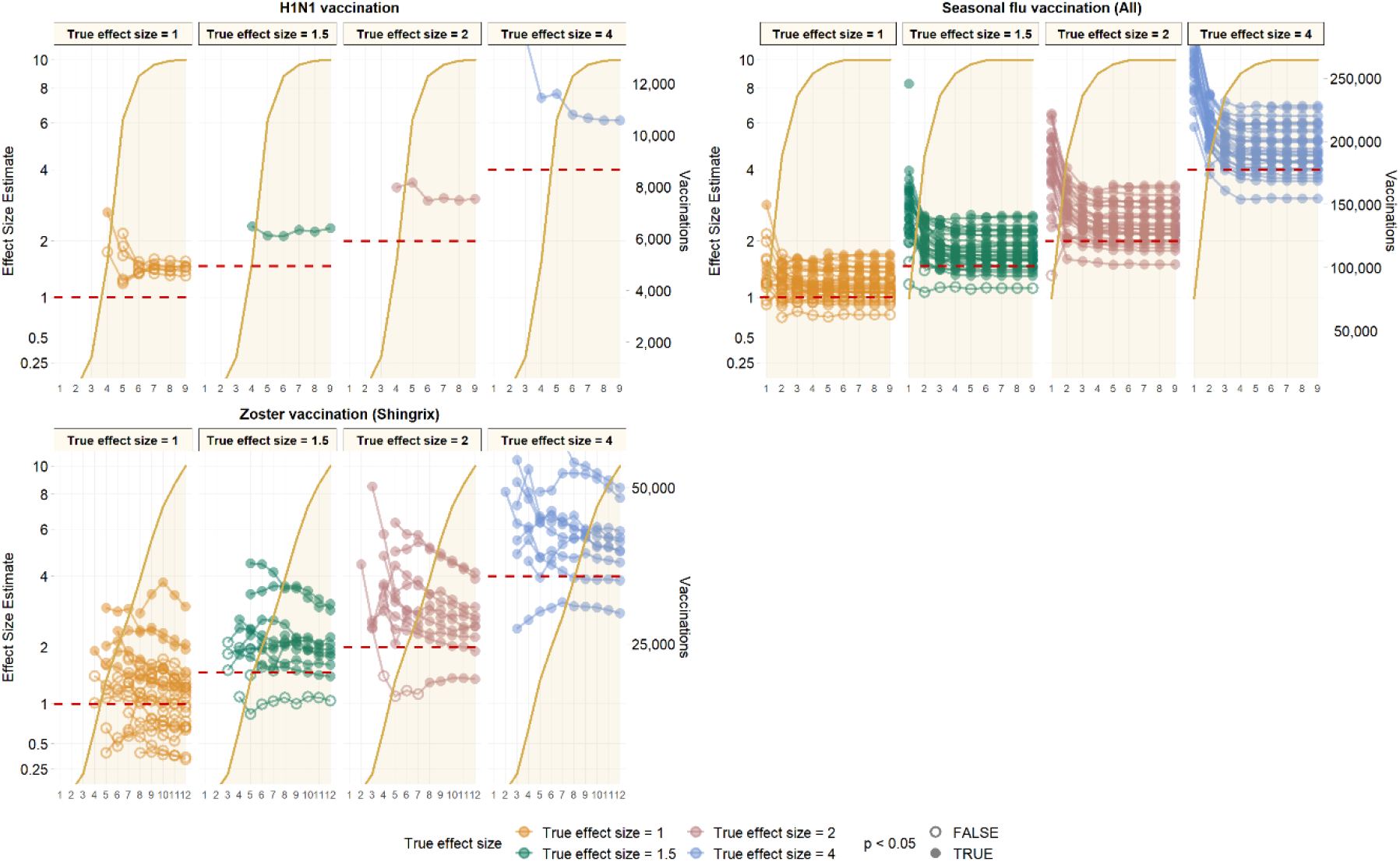
IBM MDCR (unadjusted)

**Figure S3:**
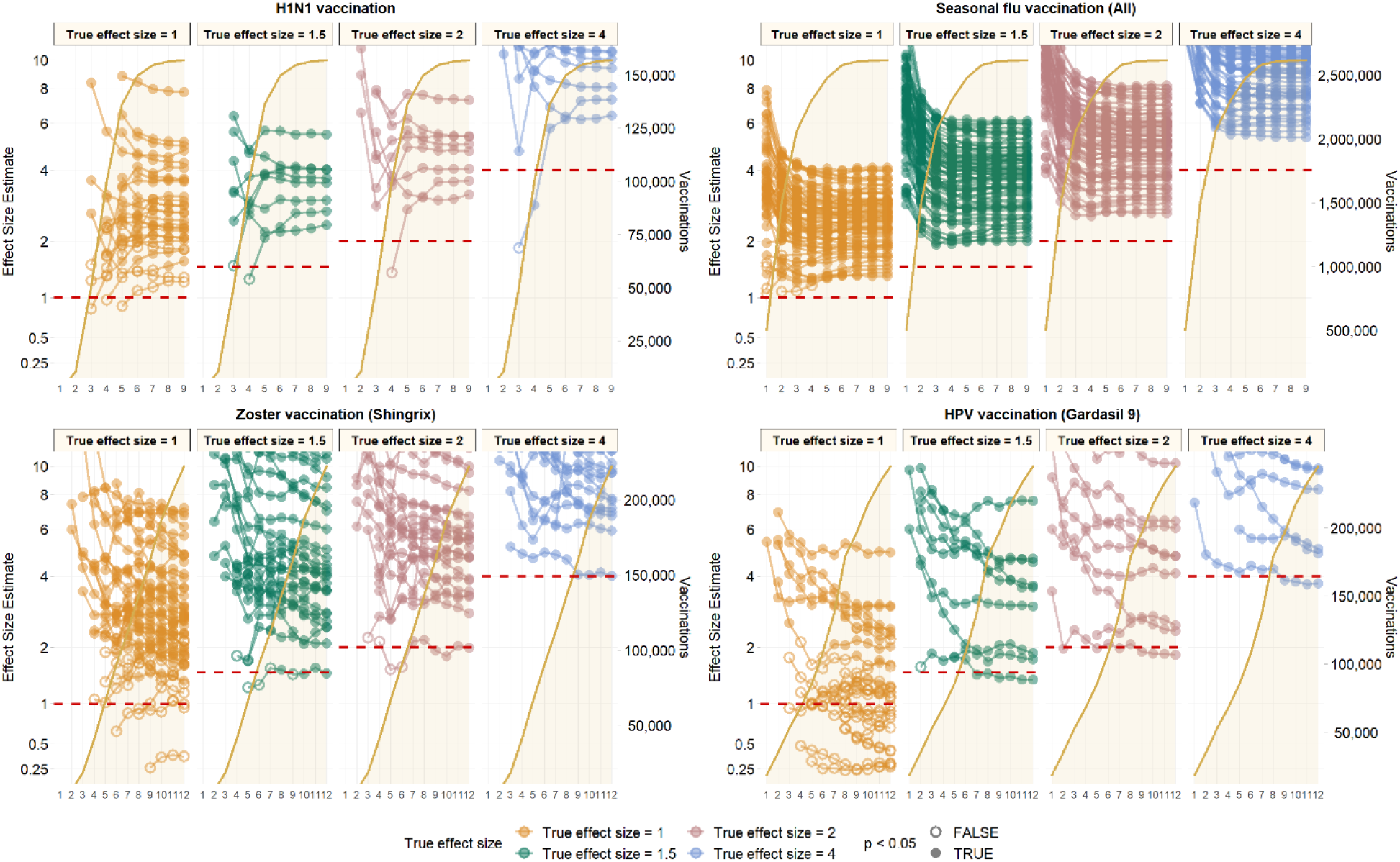
Optum EHR (unadjusted)

